# Reducing COVID-19 Airborne Transmission Risks on Public Transportation Buses: An Empirical Study on Aerosol Dispersion and Control

**DOI:** 10.1101/2021.02.25.21252220

**Authors:** Nathan J. Edwards, Rebecca Widrick, Justin Wilmes, Ben Breisch, Mike Gerschefske, Jon Sullivan, Richard Potember, Angelica Espinoza-Calvio

## Abstract

This study is one of the first COVID-19 related bus studies to fully characterize cough aerosol dispersion and control in the highly turbulent real-world environment of driving regular bus routes on both a school bus and a transit bus. While several other bus studies have been conducted, they were limited to clinical contact tracing, simulation, or partial characterization of aerosol transmission in the passenger areas with constraint conditions. When considering the risk of transmission of SARS-CoV-2 (COVID-19) and other highly infectious airborne diseases, ground based public transportation systems are high-risk environments for airborne transmission particularly since social distancing of six feet is not practical on most buses. This study demonstrates that wearing of masks reduced the overall particle count released into the bus by an average of 50% or more depending on mask quality and reduced the dispersion distance by several feet. The study also demonstrates an 84.36% reduction in aerosol particles and an 80.28% reduction in the mean aerosol residence time for some test cases. We conducted 84 experimental runs using nebulized 10% sodium chloride and a mechanical exhalation simulator that resulted in 78.3 million data points and 124 miles of on-the-road testing. Our study not only captures the dispersion patterns using 28 networked particle counters, as well as quantifies the effectiveness of using on-board fans, opening of various windows, use of face coverings or masks, and the use of the transit bus HVAC system. This work also provides empirical observations of aerosol dispersion in a real-world turbulent air environment, which are remarkably different than many existing fluid dynamics simulations, and also offers substantial discussion on the implications for inclement weather conditions, driver safety, retrofit applications to improve bus air quality, and operational considerations for public transportation organizations.

## INTRODUCTION

When considering the risk of transmission of SARS-CoV-2 (COVID-19) and other infectious diseases, ground based public transportation systems are high-risk environments without mitigations in-place. The transportation vehicles are contained air volumes where airborne transmission can be a primary mechanism, they have a large number of odd surfaces lending itself to fomite transmission due to difficulty in decontamination. In addition, social distancing of six feet is not practical on most buses and the public transportation vehicles typically have a high number of users. In a normal year, according to the U.S. National Transit Database, 9.6 billion passengers boarded public transportation buses and rail systems based on individual boardings (FTA 2020). Of concern, a recent case study has identified buses resulting in a high transmission rate, R0, where 24 of 68 people became infected after riding one a bus with one infectious person (Shen et al. 2020). Other studies describe a single person transmitted SARS-CoV-2 to 12 other individuals on two bus rides on a single day (Luo et al. 2020). In addition, there have been transmission to almost 900 workers during the pandemic within a bus transportation organization, resulting in 8 deaths (Herguth and Hurley 2020). At the time of writing this manuscript, there are 229,386 new U.S. cases of SARS-CoV-2 in a single day (JHU 2020) while several new virial mutations with higher transmission rates have emerged. Considering the possibility of close proximity to other infected passengers there is still a high potential risk of disease transmission on buses

Despite the public transportation industry’s swift response early in the pandemic to create operational guidelines that mitigate disease transmission risks there has been a large variance on implemented mitigations (Dzisi and Dei 2020; Tirachini and Cats 2020; Bushwick, Lewis, and Montañez 2020) while actionable guidance has lacked a clear basis on real world testing (NAS 2020; APTA 2020a). Fortunately, public bus transportation has had lower infection rates in comparison to their respective communities due to mitigations of mask wearing, open windows and reduced ridership with the highest percentile of COVID-19 cases from public transit systems at only 5% of the local infected population (Schwartz 2020). Nevertheless, the science and engineering community has not fully characterized the effectiveness of recommended mitigations in realistic environments while driving normal routes.

While case studies and probabilities of contact with an infected person highlight the risk of disease transmission on buses, they have not directly quantified the dynamics of airborne dispersion on buses. Many approaches in the last several years have attempted to use computational fluid dynamic simulations with ideal airflows to understand the risks and possible infectious aerosol reductions on subways (Goldbaum 2020) or other transportation environments (Hwang, DiCarlo, and Lin 2011; Löhner 2020), but they have not validated their resulting models with real world turbulent air environments in the transportation vehicles. The resulting physics simulation models are also difficult to translate into empirical field measurement and data collection. Additional research partially captures the real-world environment inside buses focusing on passenger area airflows while windows are open (Rorres 2020). Some studies have performed experiments to quantify infectious disease transmission risk on board commercial aircraft (Silcott et al. 2020; Delta Air 2020), however airflow in the aircraft passenger cabins are highly engineered to account for balanced pressures while flying at altitude. In contrast, ground transportation buses are highly turbulent with frequent stops where the doors open, passengers board or egress, windows open, seals that are not airtight, along with an ever-changing velocity and direction that affects aerosol dispersion.

Our study focuses specifically on ground transportation buses where there has been an average of 5.365 million passenger boardings on U.S. buses every day during the pandemic despite potential disease transmission risks as shown in Supplementary Information Tables S1 and S2 (APTA 2020b). In comparison to rail commuter systems or taxis, bus services are more readily accessible to people of all economic levels and available in most communities: transit buses, school buses, as well as smaller specialized transport services. With regard to the high potential risk of COVID-19 transmission on buses, broad dependency on public transportation systems, and untested assumptions in risk mitigations, our study provides complimentary yet novel test and analysis techniques to the few known comprehensive bus studies (Tawfik 2020; Zhang et al. 2021). Zhang et al. uses only two sampling instruments moved to different locations on a bus and a passenger risk metric of total number of particles at those locations, while Tawfik et al. uses three different bacteriophage virus test agents and a saturated aerosol environment to test the effectiveness of decontamination or ventilation techniques.

**Table 1:**
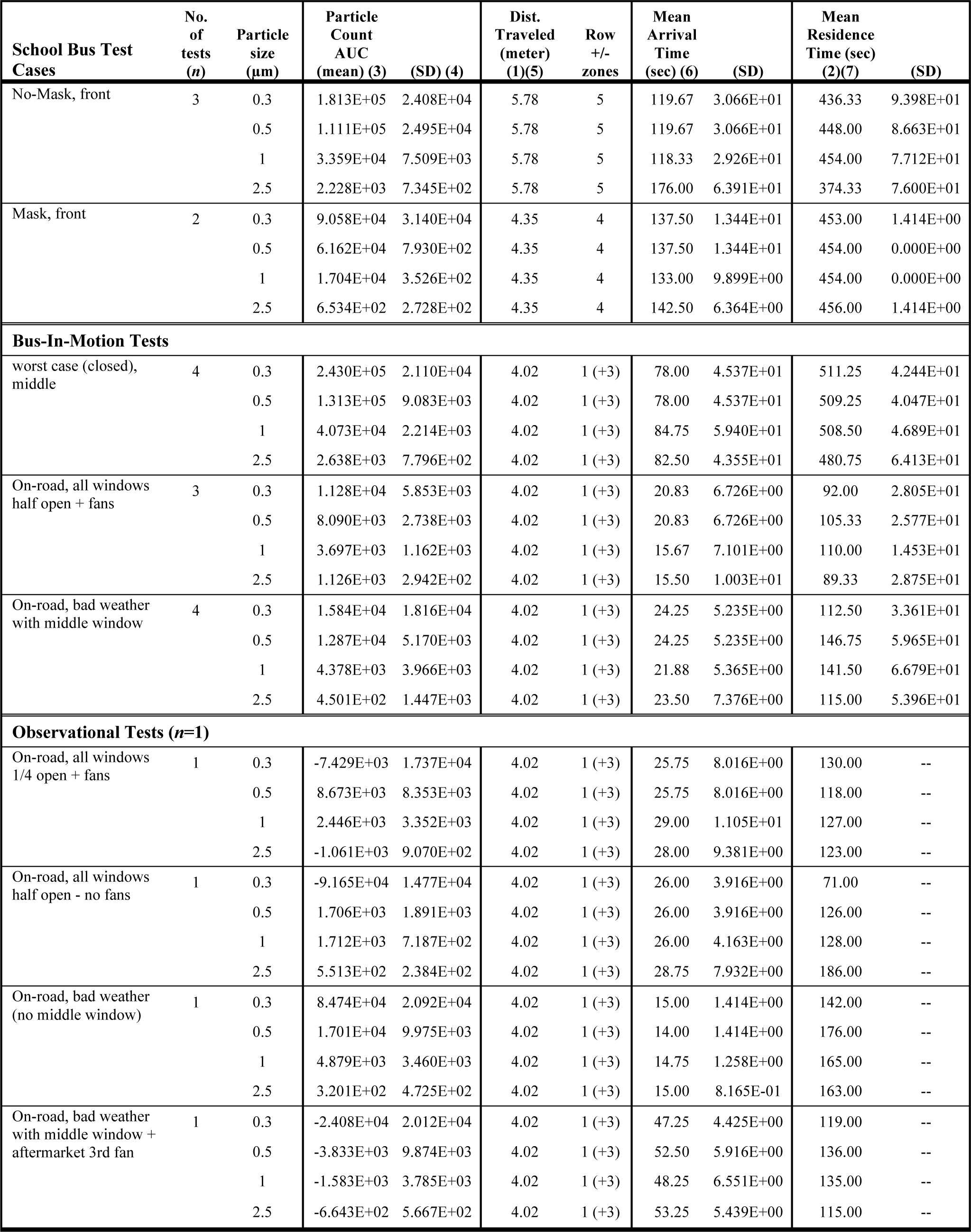
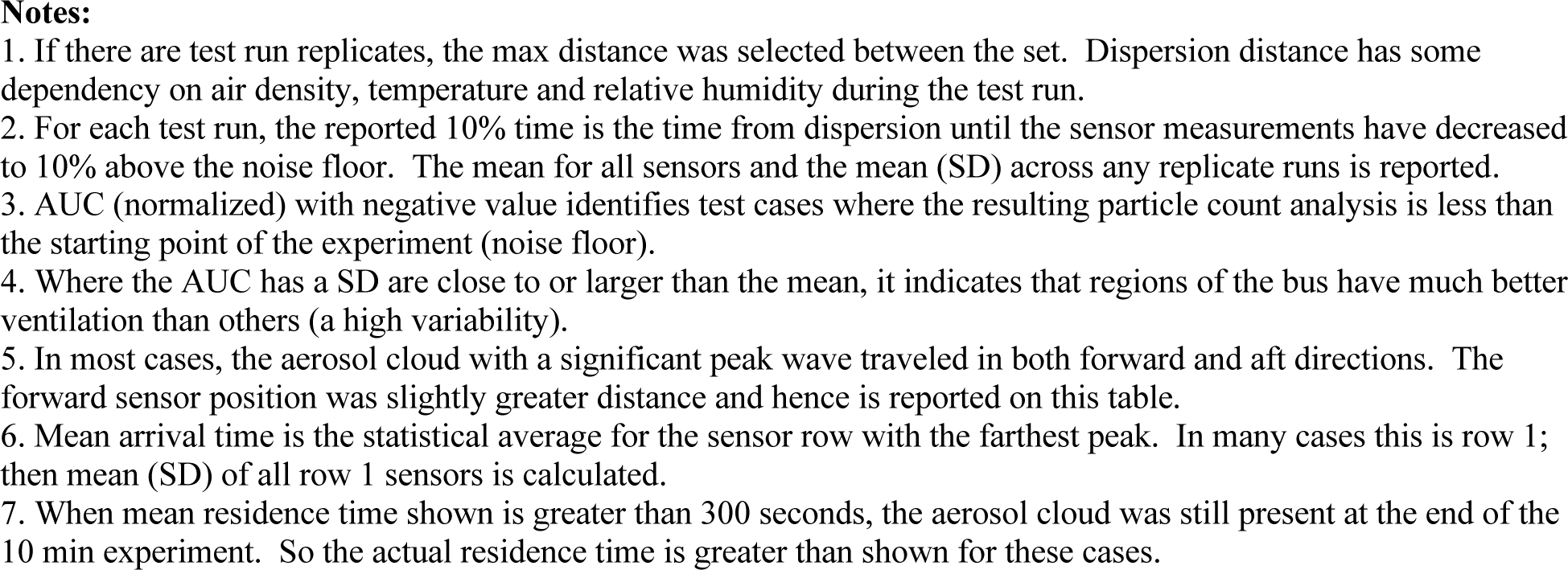
School bus aerosol dispersion and control results

| School Bus Test Cases | No. of tests (n) | Particle size (µm) | Particle Count AUC |  | Dist. Traveled (meter) (1)(5) | Row +/- zones | Mean Arrival Time |  | Mean Residence Time (sec) |  |
| --- | --- | --- | --- | --- | --- | --- | --- | --- | --- | --- |
|  |  |  | (mean) (3) | (SD) (4) |  |  | (sec) (6) | (SD) | (2)(7) | (SD) |
| No-Mask, front | 3 | 0.3 | 1.813E+05 | 2.408E+04 | 5.78 | 5 | 119.67 | 3.066E+01 | 436.33 | 9.398E+01 |
|  |  | 0.5 | 1.111E+05 | 2.495E+04 | 5.78 | 5 | 119.67 | 3.066E+01 | 448.00 | 8.663E+01 |
|  |  | 1 | 3.359E+04 | 7.509E+03 | 5.78 | 5 | 118.33 | 2.926E+01 | 454.00 | 7.712E+01 |
|  |  | 2.5 | 2.228E+03 | 7.345E+02 | 5.78 | 5 | 176.00 | 6.391E+01 | 374.33 | 7.600E+01 |
| Mask, front | 2 | 0.3 | 9.058E+04 | 3.140E+04 | 4.35 | 4 | 137.50 | 1.344E+01 | 453.00 | 1.414E+00 |
|  |  | 0.5 | 6.162E+04 | 7.930E+02 | 4.35 | 4 | 137.50 | 1.344E+01 | 454.00 | 0.000E+00 |
|  |  | 1 | 1.704E+04 | 3.526E+02 | 4.35 | 4 | 133.00 | 9.899E+00 | 454.00 | 0.000E+00 |
|  |  | 2.5 | 6.534E+02 | 2.728E+02 | 4.35 | 4 | 142.50 | 6.364E+00 | 456.00 | 1.414E+00 |
| Bus-In-Motion Tests |  |  |  |  |  |  |  |  |  |  |
| worst case (closed), middle | 4 | 0.3 | 2.430E+05 | 2.110E+04 | 4.02 | 1 (+3) | 78.00 | 4.537E+01 | 511.25 | 4.244E+01 |
|  |  | 0.5 | 1.313E+05 | 9.083E+03 | 4.02 | 1 (+3) | 78.00 | 4.537E+01 | 509.25 | 4.047E+01 |
|  |  | 1 | 4.073E+04 | 2.214E+03 | 4.02 | 1 (+3) | 84.75 | 5.940E+01 | 508.50 | 4.689E+01 |
|  |  | 2.5 | 2.638E+03 | 7.796E+02 | 4.02 | 1 (+3) | 82.50 | 4.355E+01 | 480.75 | 6.413E+01 |
| On-road, all windows half open + fans | 3 | 0.3 | 1.128E+04 | 5.853E+03 | 4.02 | 1 (+3) | 20.83 | 6.726E+00 | 92.00 | 2.805E+01 |
|  |  | 0.5 | 8.090E+03 | 2.738E+03 | 4.02 | 1 (+3) | 20.83 | 6.726E+00 | 105.33 | 2.577E+01 |
|  |  | 1 | 3.697E+03 | 1.162E+03 | 4.02 | 1 (+3) | 15.67 | 7.101E+00 | 110.00 | 1.453E+01 |
|  |  | 2.5 | 1.126E+03 | 2.942E+02 | 4.02 | 1 (+3) | 15.50 | 1.003E+01 | 89.33 | 2.875E+01 |
| On-road, bad weather with middle window | 4 | 0.3 | 1.584E+04 | 1.816E+04 | 4.02 | 1 (+3) | 24.25 | 5.235E+00 | 112.50 | 3.361E+01 |
|  |  | 0.5 | 1.287E+04 | 5.170E+03 | 4.02 | 1 (+3) | 24.25 | 5.235E+00 | 146.75 | 5.965E+01 |
|  |  | 1 | 4.378E+03 | 3.966E+03 | 4.02 | 1 (+3) | 21.88 | 5.365E+00 | 141.50 | 6.679E+01 |
|  |  | 2.5 | 4.501E+02 | 1.447E+03 | 4.02 | 1 (+3) | 23.50 | 7.376E+00 | 115.00 | 5.396E+01 |
| Observational Tests (n=1) |  |  |  |  |  |  |  |  |  |  |
| On-road, all windows 1/4 open + fans | 1 | 0.3 | -7.429E+03 | 1.737E+04 | 4.02 | 1 (+3) | 25.75 | 8.016E+00 | 130.00 | -- |
|  |  | 0.5 | 8.673E+03 | 8.353E+03 | 4.02 | 1 (+3) | 25.75 | 8.016E+00 | 118.00 | -- |
|  |  | 1 | 2.446E+03 | 3.352E+03 | 4.02 | 1 (+3) | 29.00 | 1.105E+01 | 127.00 | -- |
|  |  | 2.5 | -1.061E+03 | 9.070E+02 | 4.02 | 1 (+3) | 28.00 | 9.381E+00 | 123.00 | -- |
| On-road, all windows half open - no fans | 1 | 0.3 | -9.165E+04 | 1.477E+04 | 4.02 | 1 (+3) | 26.00 | 3.916E+00 | 71.00 | -- |
|  |  | 0.5 | 1.706E+03 | 1.891E+03 | 4.02 | 1 (+3) | 26.00 | 3.916E+00 | 126.00 | -- |
|  |  | 1 | 1.712E+03 | 7.187E+02 | 4.02 | 1 (+3) | 26.00 | 4.163E+00 | 128.00 | -- |
|  |  | 2.5 | 5.513E+02 | 2.384E+02 | 4.02 | 1 (+3) | 28.75 | 7.932E+00 | 186.00 | -- |
| On-road, bad weather (no middle window) | 1 | 0.3 | 8.474E+04 | 2.092E+04 | 4.02 | 1 (+3) | 15.00 | 1.414E+00 | 142.00 | -- |
|  |  | 0.5 | 1.701E+04 | 9.975E+03 | 4.02 | 1 (+3) | 14.00 | 1.414E+00 | 176.00 | -- |
|  |  | 1 | 4.879E+03 | 3.460E+03 | 4.02 | 1 (+3) | 14.75 | 1.258E+00 | 165.00 | -- |
|  |  | 2.5 | 3.201E+02 | 4.725E+02 | 4.02 | 1 (+3) | 15.00 | 8.165E-01 | 163.00 | -- |
| On-road, bad weather with middle window + aftermarket 3rd fan | 1 | 0.3 | -2.408E+04 | 2.012E+04 | 4.02 | 1 (+3) | 47.25 | 4.425E+00 | 119.00 | -- |
|  |  | 0.5 | -3.833E+03 | 9.874E+03 | 4.02 | 1 (+3) | 52.50 | 5.916E+00 | 136.00 | -- |
|  |  | 1 | -1.583E+03 | 3.785E+03 | 4.02 | 1 (+3) | 48.25 | 6.551E+00 | 135.00 | -- |
|  |  | 2.5 | -6.643E+02 | 5.667E+02 | 4.02 | 1 (+3) | 53.25 | 5.439E+00 | 115.00 | -- |

**Notes:**
1. If there are test run replicates, the max distance was selected between the set. Dispersion distance has some dependency on air density, temperature and relative humidity during the test run. 2. For each test run, the reported 10% time is the time from dispersion until the sensor measurements have decreased to 10% above the noise floor. The mean for all sensors and the mean (SD) across any replicate runs is reported. 3. AUC (normalized) with negative value identifies test cases where the resulting particle count analysis is less than the starting point of the experiment (noise floor). 4. Where the AUC has a SD are close to or larger than the mean, it indicates that regions of the bus have much better ventilation than others (a high variability). 5. In most cases, the aerosol cloud with a significant peak wave traveled in both forward and aft directions. The forward sensor position was slightly greater distance and hence is reported on this table. 6. Mean arrival time is the statistical average for the sensor row with the farthest peak. In many cases this is row 1; then mean (SD) of all row 1 sensors is calculated. 7. When mean residence time shown is greater than 300 seconds, the aerosol cloud was still present at the end of the 10 min experiment. So the actual residence time is greater than shown for these cases.

Our primary contributions to science are:

1. This study is one of the first to fully characterize aerosol dispersion and control on school buses and transit buses in a realistic environment of driving regular bus routes.
2. An airborne dispersion control effectiveness model that is based on passenger exposure to aerosols (time, concentration, dynamic movement) and allows for the underlying physics of an aerosol cloud and its mitigations to be more easily measured and characterized from field experimentation.
3. Empirical observations of aerosol dispersion in a turbulent air environment. The results are remarkably different than many existing fluid dynamic simulations that use ideal laminar airflows and do not account for a highly turbulent or realistic environment.
4. Offer substantial discussion on the implications of this study for inclement weather conditions, driver safety, retrofit applications to improve bus air quality, and operational considerations for public transportation organizations.

Our study not only captures aerosol dispersion patterns from a mechanical exhalation simulator, it also quantifies the effectiveness of using on-board fans, opening of various windows, use of face masks, and the use of the transit bus HVAC system while considering turbulent air and any effects of momentum inside a moving bus. The study included test runs on both school buses and low-floor transit buses where the exhalation simulator that emitted nebulized sodium chloride (NaCl) particles at airflows similar to a moderate human cough. 84 overall tests were performed using up to 28 networked high-fidelity particle counters and up to 12 airflow anemometers while the buses were stationary and while they drove normal routes. The experiments resulted in 78.3 million data points and 124 miles of on-the-road testing. Our study also analyzed the airflow data to better understand correlation to bus motion and performed computational fluid dynamics (CFD) work to validate the physics simulation model with real world data.

## MATERIALS AND METHODS

### Bus Configuration

A sixty-six seat 2013 Blue Bird Vision propane was used for the school bus experimentation. The interior configuration is very similar to many other school buses with high seat backs and slightly offset seating rows, forward and aft roof hatches, driver area, two dashboard fans, and windows that extend front to rear (SI Figure S1). The windows slide up and down and have a latch mechanism on each side to lock the windows into various notch positions. A fully open window means the top sliding portion rests on the bottom portion where only half of the window frame area is open. A half open window means that top sliding window is locked into place leaving only one quarter of the window frame open. There was no HVAC system on the school bus other than the front dashboard, two dashboard mounted fans, and two floor mounted kickspace heaters in the rear passenger area. In all on-the-road test conditions of this study, a single dashboard fan was aimed toward the central passenger aisle while the other was aimed across the windshield.

The transit bus model was a thirty-five foot, low-floor 2015 Gillig G27B model which is very similar to many other transit models where the front section of the bus is lower than the rear section to allow for easier curb-side passenger boarding (SI Figure S1). A half-height bulkhead and two small steps in the aisle was located near the rear door. The seat backs on the bus were lower than a school bus and seating configuration was more open, providing less restrictions to air movement (SI Figures S2 and S3). Only the top portion of the windows opened inward with a tilt mechanism. There were front and rear roof hatches, although access to the front hatch required use of a ladder which increases risk for drivers during normal operations and therefore was not used during the experiment. At the rear of the bus was the engine compartment which also contained the main HVAC system (Thermo King T14-M76) and ducting which had an air handling capacity of 2250 cfm controlled by the Thermo King IntelligAIRE III programmable controller.

A detailed description of experimental configurations for windows, roof hatch, fans, and HVAC system are presented in the SI Table S3. Some of the transit bus experiments used two MERV- 13 air filters (3M Filtrete 2200) with dimensions of 20 x 25 x 1 inch that were attached to the surface of the return air grille using 36 mm removable polymer tape (ShurTech FrogTape) to seal the sides of the filter to the bus and ensure all airflow goes through the filter media (SI Figure S4).

### Exhalation Simulator

The design of the mechanical exhalation simulator (SI Figure S5) is described in our prior work using a nebulized ten percent (10%) NaCl solution in distilled water to generate polydisperse particles with four general diameters of 0.3µm, 0.5µm, 1.0µm, and 2.5µm (Edwards et al. 2020). Larger particle sizes 5.0µm and 10.0µm were generated but were of insufficient quantity to accurately measure in the large air volume of the buses compared to existing dust contaminants already inside the buses. In addition, there was variability in the peak particle count generated between each test run due to the sensitivity of a NaCl recrystallization process to temperature and humidity changes throughout the day (ANSI/ASHRAE 2017).

Portable air tanks provided compressed air to the nebulizer through a low flow regulator at 103 kPa (15 psi) and also to the exhalation simulator at 827 kPa (120 psi) for both the stationary tests as well as on-the-road tests. The exhalation airflows were measured using a medical spirometer (MIR SmartOne) using the standard metrics for human respiratory function of Peak Flow (PEF) and Forced Expiratory Volume in the first second (FEV1). During this study, the simulator dispersed the NaCl aerosols with exhalation air flows of PEF (SD) of 449.8 (50.8) L/min and FEV1 (SD) of 5.80 (0.33) L which are within range of a modest cough (Lindsley et al. 2012; 2013; Hui et al. 2012; Rothenberg et al. 1987). Even though SARS-CoV-2 viral emissions and inhalation loading doses are not fully quantified by the scientific and healthcare communities, and hence a possible difference in calibrated test emissions, this method of generating aerosol particles with a simulated cough provides significant insights into aerosol dispersion patterns within the buses.

In the test cases with a mask applied, a commercial-off-the-shelf cotton fabric mask was used (ThinkDog brand from Delca Corp, item #1448827). The mask consisted of two outer layers of cotton with an inner woven layer (Delca Corp. 2020) and is expected to have a filtration efficiency of around 50 percent when compared to testing results of similar tight-weave cotton fabric and a linear increase with multiple layers (Zangmeister et al. 2020).

### Design of Experiments

Since this study did not have prior work to help shape the set of experiments, our team followed a basic exploratory workflow: 1) conduct characterization and calibration experiments to better understand aerosol dispersion in the bus environments; 2) based on observations, select a set of configurations to test with repeated experimental runs; 3) conduct face mask effect experiments with the exhalation simulator positioned at the front while the bus was static; 4) conduct the selected on-the-road experiments with repeated runs. By using this iterative approach our team was able to study the aerosol dispersion effects of each characterization experiment immediately after the run and gain invaluable insights to further shape the next experiment. However, all thirty-four of the characterization experiments are not reported in the results. The basic design of experiments for the statistical runs followed a repeating set order where the randomization in conditions was provided by the driving routes and external environments. A key benefit of the repeating set order is that it ensured the experimental runs spanned multiple temperatures and relative humidity throughout the day along with the random changes in bus direction and velocity which affected the movement of the aerosol mass (or momentum).

### Measurement and Data Collection Methods

The primary instrumentation used high fidelity particle counters (Particles Plus 8306) and anemometers (Omega HHF1001R-W) which were supplemented by GPS location data from Gauges app (version 4.0.4) on an iPhone 6S. The 8306 particle counter is a six channel particle counter that uses a NIST calibrated photometer sensor (Pariseau 2019), and was configured with a one second sampling rate in raw count mode to monitor particle diameters of 0.3µm, 0.5µm, 1.0µm, 2.5µm, 5µm, and 10µm. There were 26 particle counters placed on the school bus (Figure 1*A*), and 28 on the transit bus (Figure 1*B*) positioned approximately every other seat row to detect NaCl particles in the aerosol cloud at passenger seat positions as well as the floor and ceiling (SI Figures S2 and S3). The front most sensors were securely placed immediately behind the driver and front passenger door area. The linear distances of sensor placement are shown in SI Table S4.

**Figure 1:**
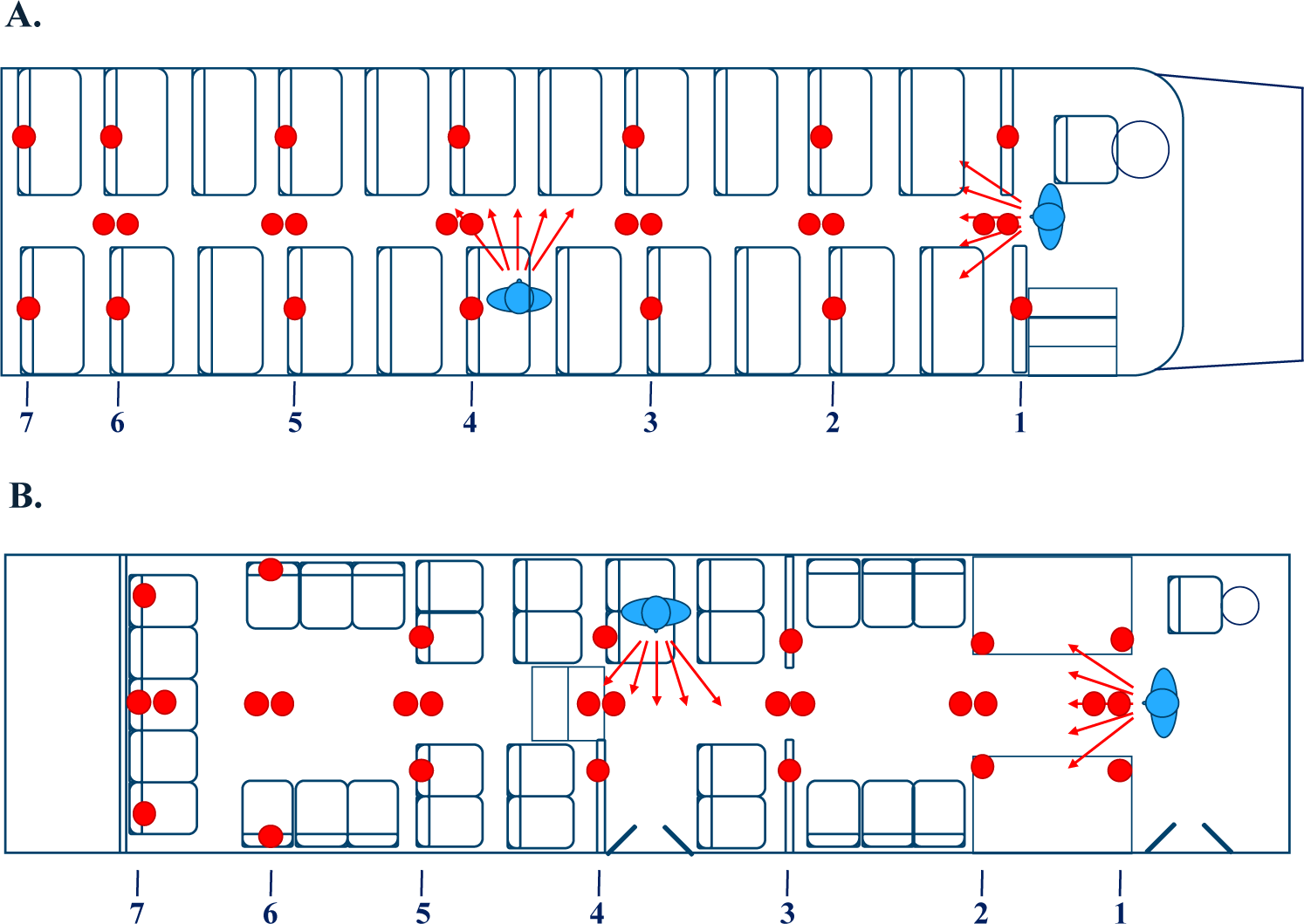
Placement of the particle counter sensors and the two simulated cough aerosol dispersion locations: (A) school bus, (B) transit bus. The dual indicators in the aisle represent the floor and ceiling mounted sensors.

Airflows on the school bus were measured with only a single anemometer due to equipment acquisition delays. The anemometer was positioned in the central aisle at an approximate head height of a standing child (118 cm from the floor), 10 cm above the level of a seat back, and provided real-time data over a USB2.0 cable interface (SI Figure S2). Twelve anemometers were used on the low floor transit bus positioned at various locations throughout the bus: forward seat areas, rear seat areas, central aisle, rear roof hatch, and the HVAC return air grille (SI Figure S3). These sensors connected to a laptop computer using their 900MHz ISM band radio communications, an USB radio receiver (Omega UWTC-REC1), and the provided Omega data recording software. All anemometers were of a hotwire type sensor to monitor single direction airflows and temperature, had measurement accuracy of 1.5% with ranges of 0 to 25.4 m/s, and were also configured to provide 1 second sampling rates.

At the beginning of each test run a timing synchronization command was sent to all particle sensors with Modbus commands over Ethernet TCP/IP and a 50 port Gigabit switch (Cisco SG250-50P-K9 PoE). Power to the Ethernet switch and exhalation simulator was provided by a 1000W portable power station (Jackery Explorer 1000). Approximately thirty seconds of data was collected from all sensors prior to aerosol dispersion which helped to establish the baseline particulate noise levels in the existing environment. During each test, the particle count data was monitored every 5 seconds with a custom supervisory control and data acquisition (SCADA) application (SI Figure S6) which provided particle count graphs to assess and observe aerosol cloud dynamics at all sensor locations. The anemometer sensor data was locally stored on the laptop computer during each experimental run. At the end of each experimental run, the whole air volume of the bus was ventilated using onboard fans and opening all windows for several minutes or using the filtered HVAC system until particulate levels subsided to nominal levels. Data was transferred from all sensors to the laptop computer using the custom SCADA software over Ethernet/Modbus.

### Safety

U.S. DOT certified cargo straps were used to secure the larger test equipment of compressed air tanks, ethernet switch, and the exhalation simulator was anchored to the steel seat structures. Sensors were secured with industrial cable ties or multiple 3M Command adhesive strips. All secured equipment was verified by the driver during a daily safety walk-through prior to on-the-road testing. Personnel safety was maintained by wearing P100 respirators while operating in close proximity to each other (due to asymptomatic COVID-19 potential) and wearing safety green, reflective traffic vests.

### Analysis Methods

Raw data files for particle count, airflow, and GPS location data were stored in CSV file format per sensor which were then preprocessed to normalize formatting and combined using Python 3.8 with the *pandas* data analysis libraries. MATLAB version R2019a was used to import the data files, compute the response variable measurements, calculate summary statistics, and plot the particle count time-series data for each sensor and experiment run. Airflow and bus velocity were also analyzed and plotted using MATLAB. Qualitative and graphical analysis was performed on the plots to validate the automated results. In some cases, the built-in *smoothdata* function was used with a Gaussian-weighted moving average filter to reduce the waveform data noise to better extract the timing measurements. In other cases, the data was too noisy for the algorithms and a manual selection of the data point was made using a graphical selection method (much like an oscilloscope trace function). Final results were imported into Microsoft Excel for formatting. The metrics and measurements used for comparison of effectiveness are presented in the next section.

The 3-dimensional CAD model computational fluid dynamics (CFD) simulation was developed using CREO 4.0 from PTC. The CFD analysis and flow field visualization used an Ansys 2020 R2 CFX high performance computing (HPC) license to run on 16 multiprocessor cores along with a *cfd_solve_level2* license for the multiphase flow solver.

### Airborne Dispersion Control Effectiveness Model

In order to understand the results of this study we must first explain the novel airborne dispersion control effectiveness model used to analyze the data and present the results of risk and control. While there are underlying physics as the basis for aerosol cloud dispersion, its direct measurement and associated mathematics are not as easily translatable to risk reductions that can be applied by broad non-scientific communities in public transportation. The proposed airborne dispersion control effectiveness model is based on passenger exposure to aerosols (time, concentration, dynamic movement) and allows for aerosol cloud dynamics and effectiveness of mitigations to be more easily measured during field experimentation. Our effectiveness model shown in Equation 1 is used to quantify the effectiveness of each scenario tested on the dispersion of specific particle diameters *d* (0.3µm, 0.5µm, 1.0µm, 2.5µm) and is a function of four primary factors:

Where:

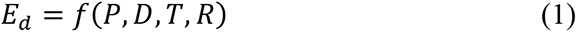

*P* = overall particle count characteristics of the aerosol cloud dispersion.

*D* = dispersion pattern of the aerosol cloud (distance traveled, direction).

*T* = timing characteristics of the aerosol cloud (arrival time).

*R* = residence of particles (or residual amount).

Most of the common guidance for reducing airborne infectious disease transmission is based primarily on the factor *R,* residence of particles (Mittal, Meneveau, and Wu 2020; Sze To and Chao 2010; Riley, Murphy, and Riley 1978). However, in empirical observation of aerosol cloud dispersion, a multivariate model is more holistic and representative of real-world disease transmission considering the dynamics of people, their movement behaviors, their respirations, and turbulent air environments. While the theory and detailed basis for the effectiveness model will be described in future work, we present the results of our study from measurements of these four factors individually which are distinctly visible in the time-series particle count waveforms (SI Figures 7, 8, 9, 10).

### Particle Count (P)

The overall particle count characteristics of the aerosol clouds in this study are quantified by a summation of particle count measurements over a ten-minute period at each sensor, known as the area under the curve (AUC). We selected the AUC of particle count rather than of particle concentration as it better suited to measuring the overall characteristics of an aerosol cloud (incline, peaks, decline, anomalies from turbulence, etc.) and is more closely related to infectious disease dose-response research which uses the number of virions in an aerosol for a given period of time. The AUC used in this study was normalized by subtracting the noise floor from raw AUC values (Figure 2*A*) so that it is indicative of the NaCl aerosols introduced by the experiment at the current temperature and relative humidity within the buses. The noise floor was determined using an average particle count immediately preceding the dispersion event. A lower or negative AUC equals good air exchange or filtration where the particle count is less than at the start of the experiment run. A higher value for AUC indicates less effectiveness at reducing particle counts over time.

**Figure 2:**
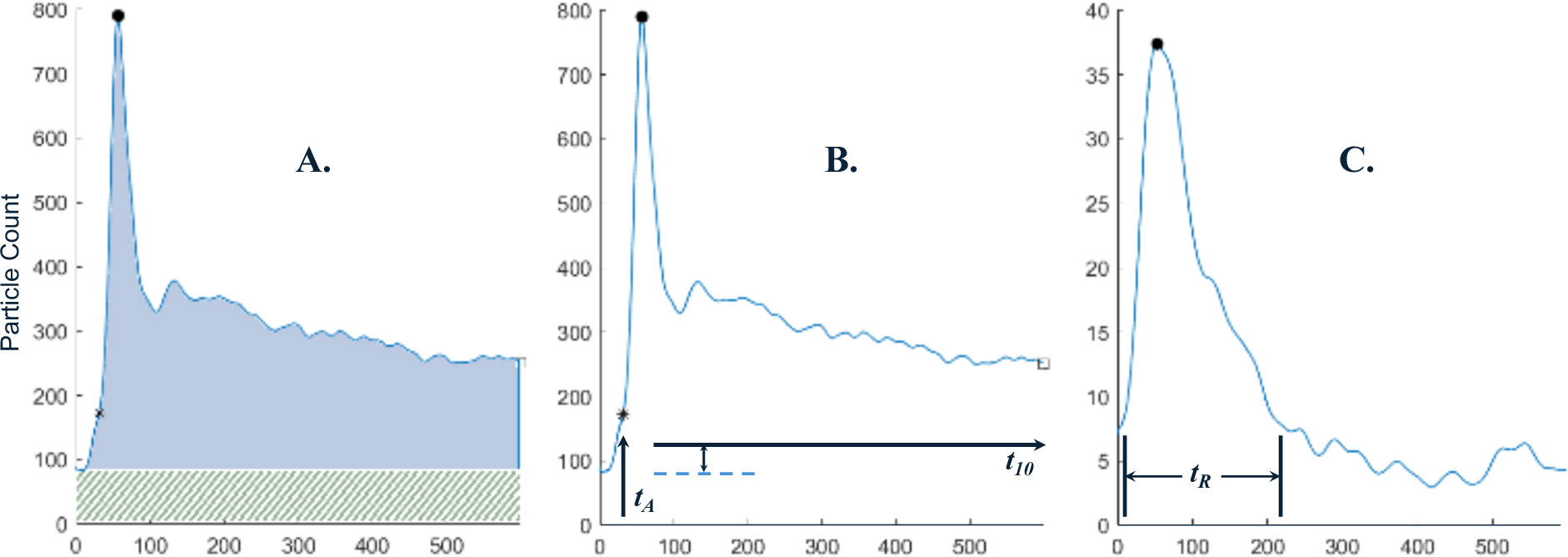
Examples of the measurements performed on the time-series aerosol particle data for a distinguishable aerosol waveform: (A) particle count AUC excluding the noise floor, (B) arrival time, *tA*, also showing that a 10% threshold time, *t_10_*, is never reached after dispersion, and (C) aerosol residence time, *t_R_*, at 10% above the noise floor.

### Dispersion Pattern (D)

The dispersion pattern of the aerosol cloud is measured by linear distance and direction that it aerosolized particle cloud travels from the dispersion location to sensors where the peak particle count has a distinguishable wave (i.e. particle count waveform with significant slope increase). At locations beyond this distinguishable wave, aerosol particles appear to gradually increase overtime as the concentration equalizes throughout the environment. Distance traveled is measured by the linear distance from the exhalation simulator to the distal sensors in units of meters and instrumentation rows (SI Table S4). Less distance traveled or a directional vector away from passenger seating both indicate better dispersion control.

### Timing Characteristics (T)

The timing characteristics of airborne particle dispersion relate to a person’s potential exposure time to an aerosolized cloud containing infectious disease, extended time in the aerosol increases the probability of inhalation and disease transmission. The timing characteristics in this study are presented as the mean arrival time of an aerosol cloud (Figure 2*B*) at the farthest location of a distinguishable wave. Arrival time of the aerosol cloud indicates speed of spread and is also compared across multiple experiment runs to observe any airflows from windows or HVAC systems that compete with the aerosol cloud dispersion. A slower arrival time is generally better as it gives people time to react and avoid an infectious aerosol cloud from a sneeze or cough.

### Residence of Particles (R)

The residence time of particles is the average time after an aerosol cloud arrives for the particle count measurements to decline to nominal levels (Figure 2*C*). The value of 10% above the noise floor was chosen as a nominal level threshold as most face coverings or fabric masks offer protection at this minimal level. For this study we report the residence time at the farthest noted sensors as an estimator for a worst-case scenario.

## RESULTS

### Effectiveness of Face Masks on Buses

With a commercially available cotton face mask applied to the exhalation simulator at the front of each bus, it reduced the overall particles counts, reduced the distance of the peak aerosol cloud travel, and increased the aerosol cloud arrival time; all of which reduce passenger risk of potential exposure to airborne contaminants. The results are presented for school buses (Table 1), for transit buses (Table 2), and summarized with overall percentile changes (Table 3). A mask applied on the school bus resulted in a mean decrease in particle count AUC and mean increase in aerosol cloud travel time across the four particle diameters (0.3µm, 0.5µm, 1.0µm, 2.5µm). The longest distance traveled by the aerosol cloud on the school bus was reduced only by 1.43 meters although the overall velocity of the aerosol cloud decreased by 27.27% from detailed analysis. Additionally, the particle count waveform was significantly attenuated by an average of 53.08% for the smaller three particle diameters (SI Figure S7). Likewise, application of a face mask on the transit bus resulted in a mean decrease in particle count AUC and a mean decrease in aerosol cloud travel distance by 4.77 meters and an associated faster arrival time due to the reduced distance. The overall velocity of the aerosol cloud on the transit bus decreased by 25.75%. In contrast, a mask increased the residence time of the aerosol particles across the four particle diameters for both the school bus and transit bus despite the reductions in overall particles.

**Table 2:**
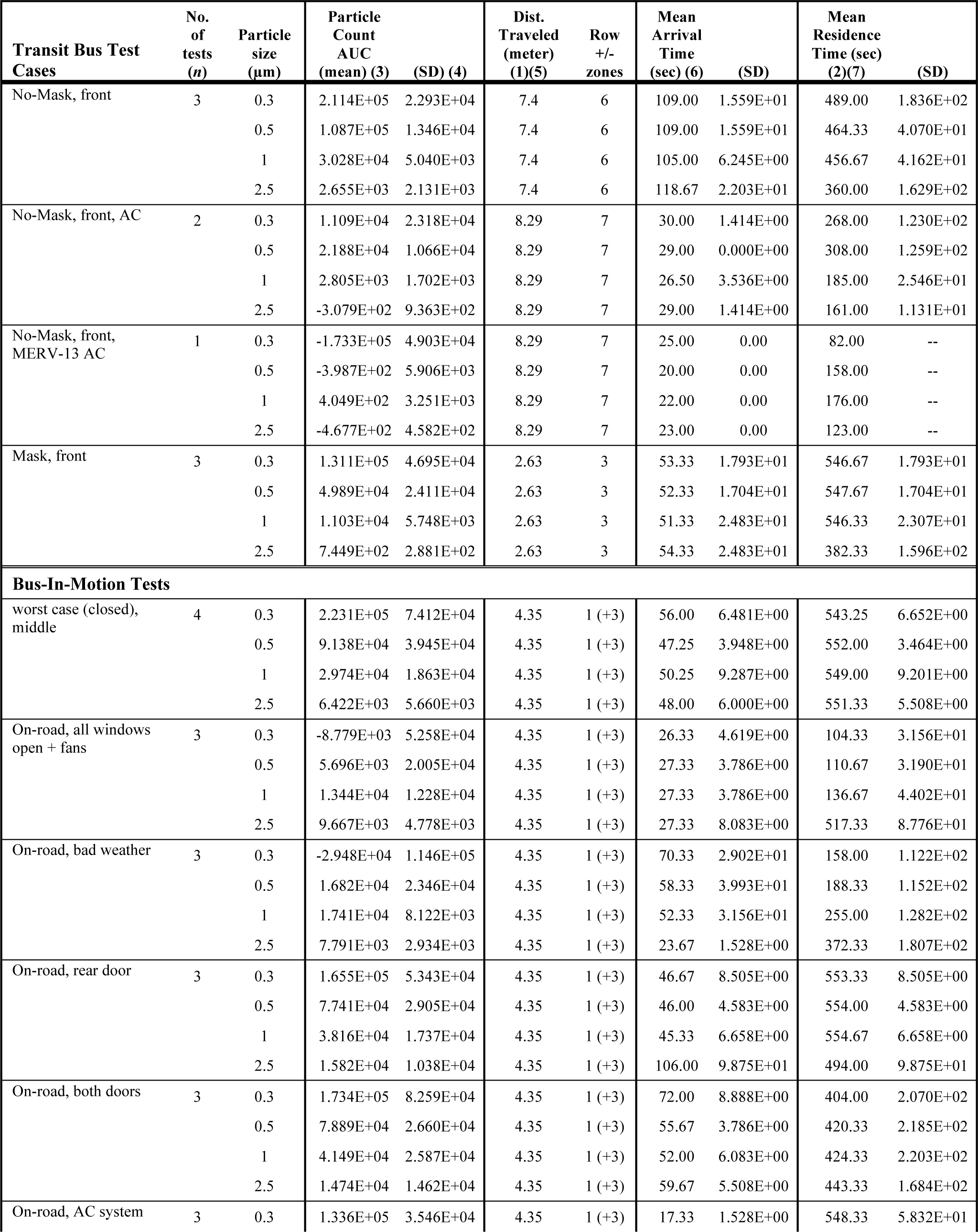
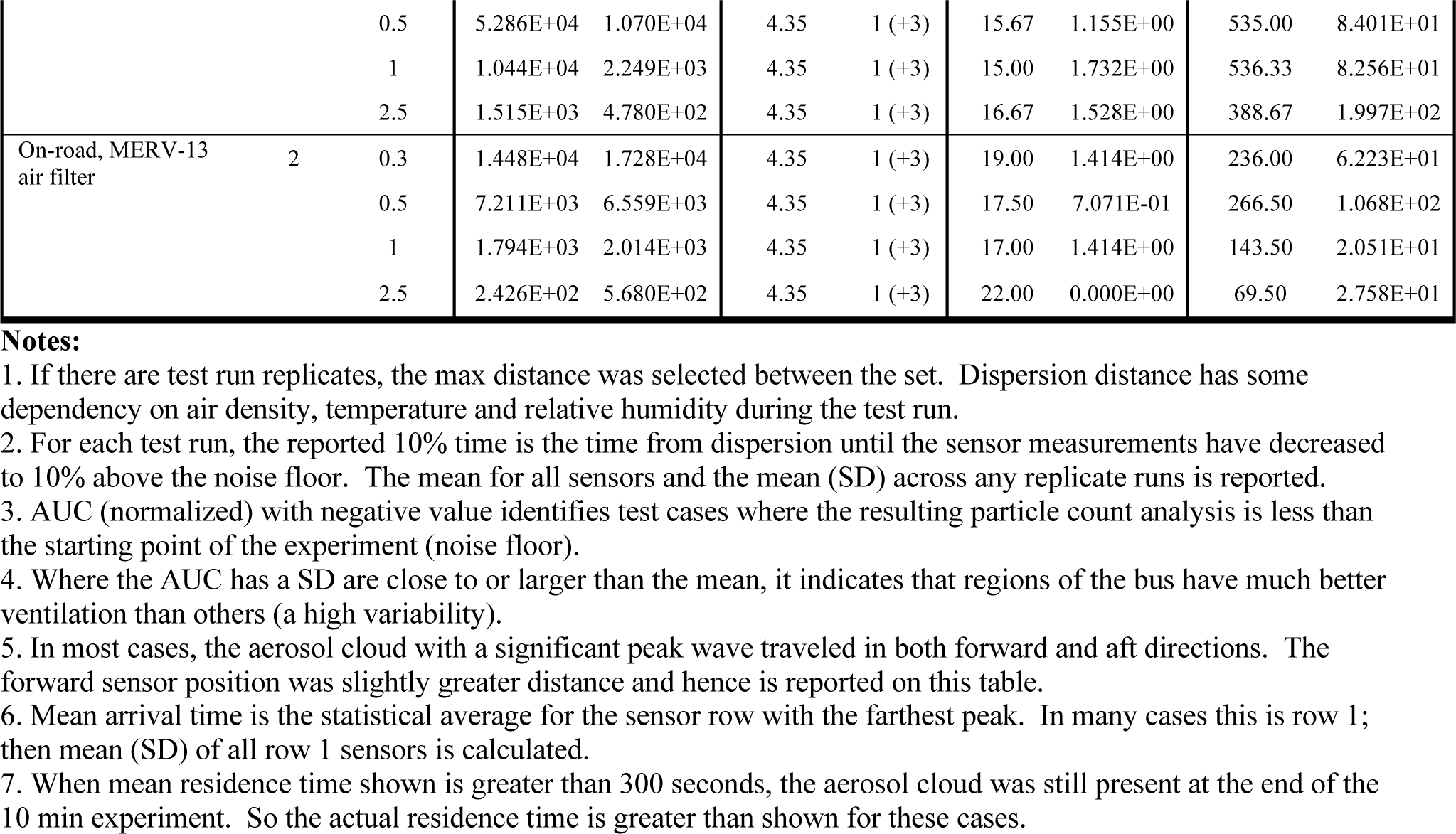
Transit bus aerosol dispersion and control results

### Effectiveness of Opening Windows, Roof Hatches, and Dashboard Fans

In general, we summarize the ventilation results into two categories: A) all windows and easily accessible roof hatches open; and B) a few select windows opened to limit exposure to the external environment for an inclement weather configuration. The optimal air exchange configuration of all windows and accessible roof hatches along with dashboard fans resulted had significant reductions in the overall particle count AUC, an average of 84% on school buses and 50% on transit buses, and also reduced the mean residence time of the aerosol particles by 80% in school buses and 60% on transit buses (Tables 1 and 2, SI Figure S8). All of these results indicate a reduced passenger risk of potential exposure to airborne contaminants. However, the peak aerosol cloud traveled the maximum distances from the dispersion location, spreading throughout the bus three rows forward and aft. The experiments with windows, roof hatches and fans also resulted in decreased aerosol cloud arrival time (or faster speed of travel) due to the significant airflow throughout.

The reduced opening window configuration for inclement weather also resulted in reductions of particle count AUC and mean residence time of the aerosol particles (SI Figure S9). The distance of the peak aerosol cloud travel stayed the same at the maximum distances from the dispersion location (both forward and aft). The aerosol cloud arrival time was slower in comparison to all windows open yet faster than no windows open on the school bus and slower than no windows open on the transit bus, highlighting the key differences in air volume and seat configurations.

### Effectiveness of Opening Transit Bus Doors

The results of opening the transit bus doors at all bus stops along a route are shown in Table 2 for tests with rear-door only and both rear and front doors. In both test conditions all windows were closed, and the rear roof hatch was opened to allow a pressure relief inflow and avoid stagnation with the air volume in the rear section of the bus.

With the rear door open at all bus stops, the particle count AUC had some reductions for particle diameters of 0.3µm and 0.5µm but increases for diameters of 1.0µm, 2.5µm due to external environment contaminants such as dust or smaller micronic pollen at the bus stops (Agarwal et al. 1984; Takahashi et al. 2003). There was no difference observed in the aerosol cloud travel distance compared to the worst case with all windows closed (Figure 3*A*), and negligible changes to the mean residence time. The mean arrival time of the aerosol cloud decreased with the smaller three particle diameters at the forward most sensor row, yet resulted in a significant slowdown with the 2.5µm particle diameters. When analyzing the particle count waveforms of the aerosol cloud motion, localized perturbations were highly visible at sensors near the rear door but negligible at other locations.

**Figure 3:**
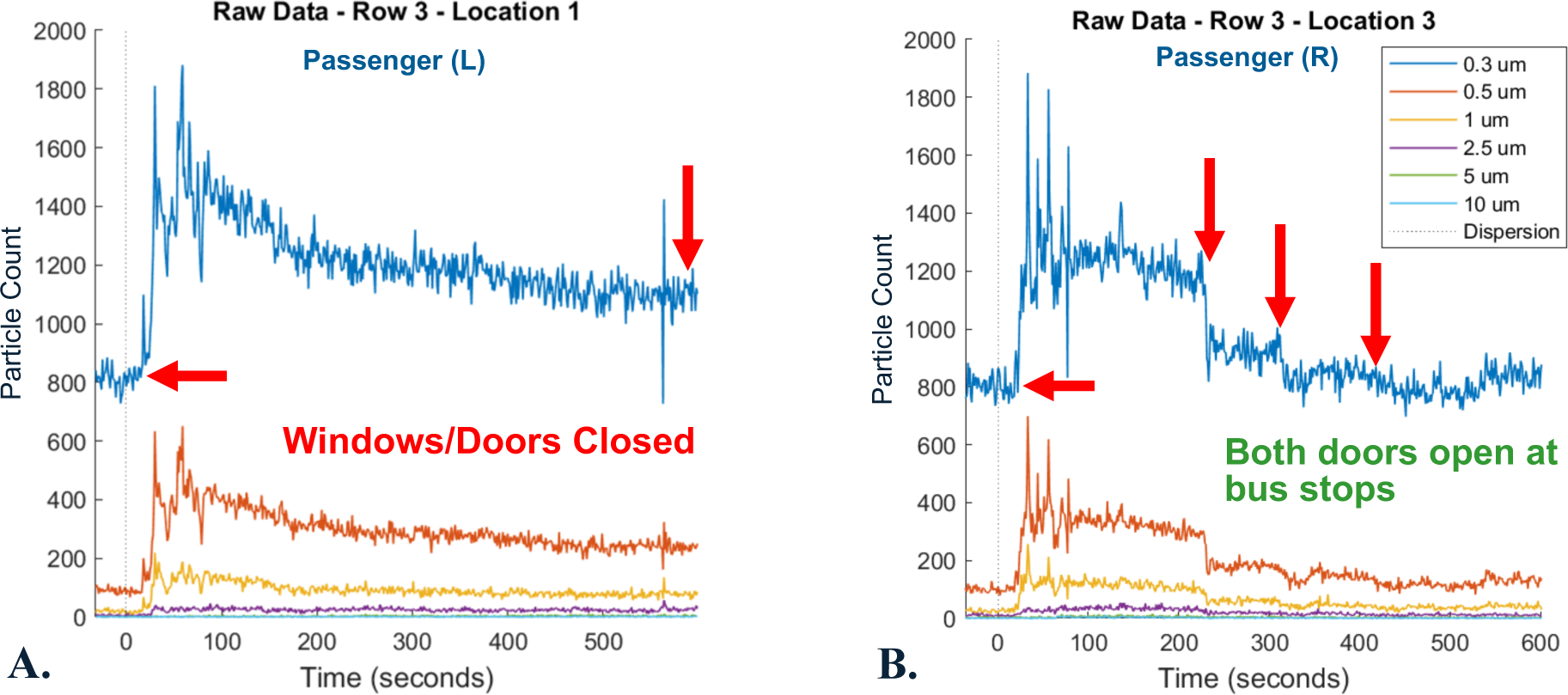
Particle count time-series waveforms from transit bus test with arrows indicating the baseline noise floor (left) and any decreases in aerosol residence (right). (A) shows sensor data from single location with all windows closed. (B) Shows a step function decrease effect when both doors were opened at bus stops.

With both doors open at all bus stops, the particle count AUC also had some reductions for particle diameters of 0.3µm and 0.5µm and increases for diameters of 1.0µm, 2.5µm due to external environment contaminants. This test configuration also resulted in no differences to the aerosol cloud travel distance compared to the worst case with all windows closed. The aerosol cloud arrival times slightly increased for all particle diameters due to the increased airflow, and the mean residence time decreased by 19.59% to 25.63% depending on the particle diameter. Overall, having both the front and rear doors open at all bus stops along a route had better performance at reducing particle exposure risks than only a single door open with highly visible changes in the particle count waveform for some test runs (Figure 3*B*).

### Effectiveness of Transit Bus HVAC System and MERV-13 Air Filter Modification

To compare and contrast effects of the HVAC system and air filter modifications, there were two primary test conditions: the AC system on; and two MERV-13 air filters covering the HVAC return air vent. Both conditions were tested with two different dispersion locations: one at the front of the stationary bus with the exhalation direction toward the rear; and the second in the middle of the bus during on-the-road testing.

The effect of the AC system without an air filter resulted in a significant decrease in the particle count AUC and rapid increase in the mean arrival time (aerosol velocity) reaching the distal locations. The mean residence time of the aerosol particles decreased some, but it was observed that reductions in aerosol particles after the peak dispersion were gradual. Additionally, the nominal threshold level of 10% above the noise floor was never reached by the end of the ten-minute time period in a number of experimental runs (SI Figure S10*A*).

By adding the retrofit MERV-13 air filters to the HVAC return air vent in the back of the transit bus, the effectiveness of removing aerosol particles increased significantly. The resulting particle count AUC with the air filters resulted in an average of 93.95% improvement with aerosols dispersed from a middle location during bus in-motion testing (Table 3). The other key improvement was the aerosol mean residence time which had a maximum of 266.5 seconds (or 4.44 minutes), but in most cases the residence time was less than 176 seconds or 2.93 minutes (SI Figure S10*B*). Similar to the previous HVAC tests, adding MERV-13 air filters also had increases in the mean arrival time at the distal locations (aerosol velocity).

**Table 3:**
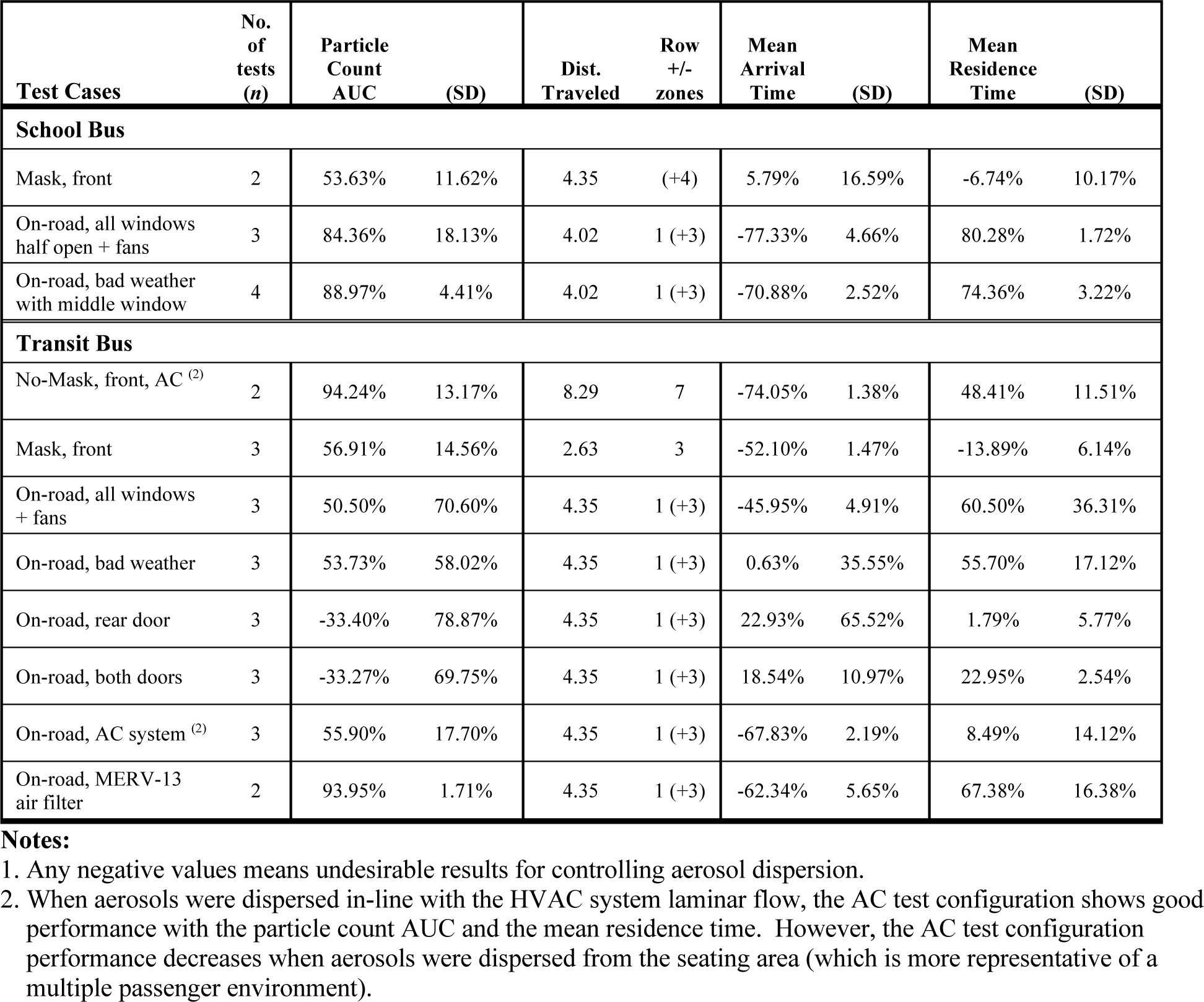
Percentile improvements compared to the worst-case scenarios (no-mask, windows closed). Average across all particle diameters.

### Preliminary Airflow Analysis

While the primary focus of this study is aerosol dispersion and identifying the optimal risk reducing controls, we present preliminary comparison results of passenger area airflows to overall bus velocity while driving the various routes. When interpreting the results, it is important to understand the main airflow effects are from the bus velocity of driving forward. However, the internal airflows are also affected by external wind or crosswinds that might blow through the open windows which can help rapidly ventilate the aerosol particles.

With all school bus windows half open, the airflow appears to have a horizontal asymptote nearing 1.5 m/s as the maximum airflow in the central aisle (SI Figure S11*A*). Likewise, the horizontal asymptote and maximum central airflow for the front, middle, and rear window inclement weather configuration is around 1.0 m/s (SI Figure S11*B*). A single observational test run with all windows one-quarter open (2 notches down) resulted in a slightly higher maximum airspeed around 1.1 m/s in the central passenger aisle (SI Figure S11*C*). The fourth configuration of the frontmost and rearmost windows open has the least airflow response with the majority of measurements less than 0.75 m/s (SI Figure S11*D*) and appears to be more susceptible to poor airflow phenomena with the bus velocity around 12 m/s and 17 m/s.

On-the-road experiments with all the transit bus windows and rear roof hatch open there was modest airflow in the seat areas with a maximum airflow measurement of 1.25 m/s (SI Figure S12*A*). The average linear fit of airflow ranged from 0.14 m/s at slow bus velocities up to approximately 0.4 m/s at the higher bus velocities. In comparison, there was a maximum airflow measurement of 0.8 m/s with only front and rear windows and roof hatch open (SI Figure S12*B*), and much less average airflow of 0.05 m/s at slow bus velocities up to 0.2 m/s at the higher bus velocities. In both cases the rear elevated seat area had slightly more airflow than the forward low-floor seats indicating that natural ventilation had more effect on the elevated rear seat areas. However, further analysis is needed to correlate airflows with aerosol dispersion and control in the front and rear sections.

The results of the AC system running with all the windows closed resulted in higher steady airflows in the passenger area at an average of 0.55 m/s in the forward seating area, 0.36 m/s in the rear seating area, while the HVAC return air maintained an overall airflow of 2.5 m/s (SI Figure S12*C*). With the retrofit MERV-13 air filter applied to the return air grille, airflows were slightly less with an average of 0.41 at m/s at the forward seating area, 0.3 m/s at the rear seating area, and 2.0 m/s at the return air grille (SI Figure S12*D*). In both cases with the HVAC system, the forward seating area maintained higher steady airflow than the rear elevated area.

### Computational Fluid Dynamics

Full results of validating our computational fluid dynamics (CFD) model with the field test data is not presented in this work, however basic visualization from the CFD analysis is useful to understand the bus passenger indoor air environments. The CFD simulation developed from empirical field test data is that the low-floor transit bus has an extremely turbulent flow field which characterizes the complexity of this environment (Figure 4). An important observation from analyzing field experiments of the school bus is that the high seat backs and seating configuration create a baffle like effect and cause the aerosol dispersion jet to diffuse more quickly. CFD analysis of the school bus provides a visualization of this effect where regions of high aerosol concentration are present at the front side of seats, highly visible at *t* = 60 seconds and *t* = 120 seconds (SI Figure S13).

**Figure 4:**
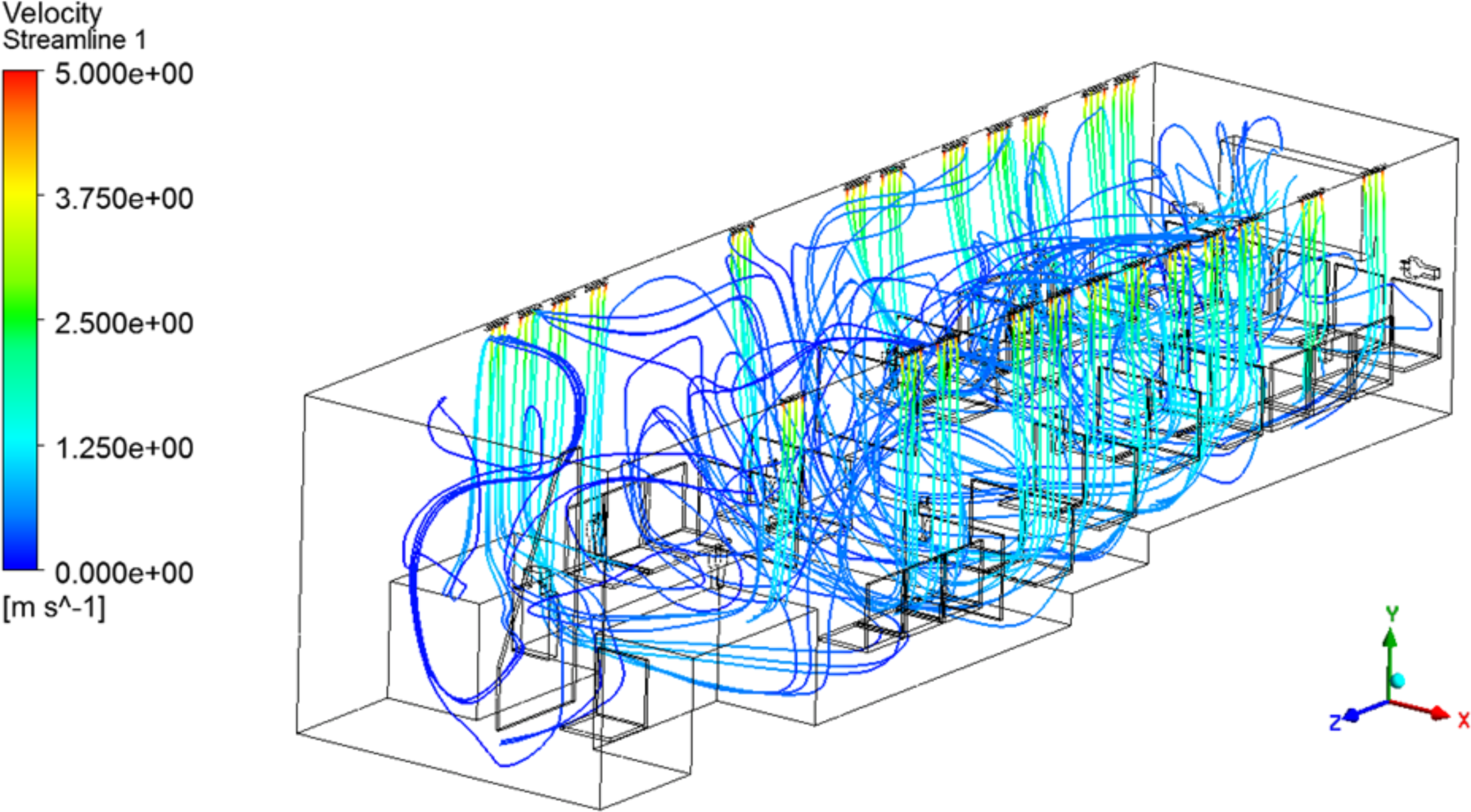
Airflow streamlines from CFD analysis of the low-floor transit bus showing an extremely turbulent air environment. Input parameters and initial airflow measurements were obtained from field test data.

### Results from Other Observational Tests

Several other experimental tests were conducted during this study and are presented in Tables 1, 2, and 3 as single run observational tests only. Caution must be applied when inferring conclusions from these individual test runs given the complex air environment inside the buses.

However, when coupled with the real time monitoring of particle count waveforms, they helped to shape the final repeated experimental runs (an *n* greater than one) for the statistical analysis.

## DISCUSSION

With 84 overall experiments (34 characterization tests and 50 statistical runs), this study demonstrates not only the complexity of the turbulent air environment in a moving bus, but also clearly shows that air exchange, air flow, filtration, and masks have a significant effect on aerosol dispersion control. One important conclusion is that the wearing of a mask by an infected person in the shared airspace will directly affect the factors of *P*, *D*, *T* and *R* and significantly reduce overall airborne dispersion in a bus. Wearing of masks reduced the overall particle count released into the bus by an average of 50% or more depending on mask quality and reduced the dispersion distance by several feet. When masks were not worn, dispersed particles spread rapidly throughout the whole bus. Another important conclusion from the results is that maintaining airflow, fully or partially, reduces aerosol particle counts and the associated risks.

The results of this study also identify tension between different metrics of reducing passenger exposure risk to aerosols. For example, in one test condition particle count AUC improves drastically from effective ventilation, but aerosol cloud arrival time simultaneously shows an a slightly undesirable result – faster speed of dispersion toward other passenger areas. The example also highlights that risk-based mitigations may not have a single optimal solution for all desirable effects and a risk-gradient approach must be used. In other words, it is important to consider the tradeoffs between each desired outcome measurement and its relationship to infectious disease risk reductions even if the aerosol transmission has not been fully quantified by the health science community.

### Observations of Aerosol Cloud Behavior

Throughout the experiments, we observed high air turbulence within the buses and peculiar aerosol cloud motion that could have an impact on passenger exposure. The first observation was that an aerosol cloud was subject to momentum. With drastic changes in bus velocity and direction, the particle count waveforms decreased as the aerosol cloud moved away from some locations while increasing at other locations and appeared to follow Newton’s physics laws of motion during the initial changes by the bus. Considering that the net total of aerosol NaCl particles did not change during the tests with all windows closed, this observation provided a method for identifying aerosol motion characteristics.

Secondly, during some of the experiments we observed backflow behavior of the aerosol cloud, where the cloud motion originated from the back of the bus and increased particle counts moving forward. This behavior led to the opening of a middle window open on the school bus for a bad weather configuration, which provided some airflow relief and reduced the backflow effects.

A third observation is that the back row of seats tended to accumulate more particles (windows closed configuration) and should be considered as locations of higher exposure risk. To summarize, motion or dispersion of a potentially infectious aerosol cloud on a bus may unexpectedly expose passengers at distant locations on the bus; this also emphasizes the importance of passengers wearing face masks for protection.

We also observed interesting aerosol cloud behavior from the effects of both transit bus passenger doors open at all bus stops. During some of the experiments we observed a significant reduction in aerosol particles with every door opening event, where the particle count waveform appeared as a decreasing step function when the doors were opened (Figure 3*B*). This was not always the case in other experimental runs and appeared to have some dependency on the external environment (e.g. wind direction, etc.) at each bus stop. However, the important concept is that with both doors open there is a greater opportunity for fresh air exchange of the potentially contaminated air inside the bus.

A final observation is that the transit bus rear door bulkhead and steps leading to the raised rear seating area help diffuse an aerosol cloud emitted at the front of the bus and traveling toward the rear. When the exhalation simulator was placed in the middle aisle near the driver’s area, the time-series particle count waveforms had a highly distinguishable peak waveform for the first three sensor rows before the bulkhead and steps. Any waveform from sensors behind the steps or bulkhead displayed an attenuation of the peak and more of a gradual increase in particle counts. This observation indicates that other bulk object barriers in the bus are likely to have similar effects of diffusion and slowing of the aerosol cloud.

### Implications for Inclement Weather

The challenges presented by inclement weather and potential closing of windows highlights the crux of an imperfect risk-based problem. In some geographic regions, weather conditions can drastically change throughout any given day which may require windows to be closed to prevent water, snow, high humidity, or severe dust of a windstorm from entering the passenger or driver areas. However, while closing windows may increase passenger comfort levels during inclement weather it also increases passenger risk of exposure to potentially infectious aerosols from the reduced airflow and air exchange. Passenger safety is paramount in public transportation, so the challenge is to balance risks with passenger comfort levels.

One initial concern is the potential wind chill effect on passengers from bus windows being opened during inclement weather. According to the National Oceanic and Atmospheric Administration, wind chill is only defined for wind speeds or airflows above 1.34 m/s (3 mph) (NOAA 2001). As previously discussed in the preliminary airflow analysis, the maximum observed airflows with all windows open were 1.5 m/s on the school bus (SI Figure S11*A*) and1.25 m/s on the transit bus (SI Figure S12*A*). The inclement weather window configurations resulted in 1.0 m/s school bus (SI Figure S11*B*) and 0.8 m/s transit bus (SI Figure S12*B*). Similarly, another airflow study was conducted on transit buses with comparable results (Rorres 2020). Conclusively, wind chill may only apply to a few select seat areas in the direct jet stream from an open window.

Another concern is the impact that low human comfort levels may have on ridership with windows open during inclement weather, hence a comparison with indoor HVAC design guides is conducted to estimate comfort levels. The American Society of Heating, Refrigeration, and Air-Conditioning Engineers (ASHRAE) found that many people are comfortable where the effective draft temperature is between -3 °F and +2 °F with air speeds less than or equal to 0.35 m/s (Arens et al. 2015). Likewise, the U.S. Bureau of Reclamation’s design guide for HVAC systems recommends air motion from 0.25 m/s (50 ft/min) to 0.38 m/s (75 ft/min) for sedentary work, and from 0.51 m/s (100 ft/min) to 1.52 m/s (300 ft/min) for high activity or maintenance work (USBR 2006). Fundamentally it can be inferred that the inclement weather window configuration on buses may be slightly uncomfortable in cold weather due to air motion but still within a reasonable range for human comfort.

Inclement weather conditions were taken into account with the “bad weather” window configurations (SI Table S3) which have the front, middle, rear windows open (school bus) or the front and rear windows (transit bus) which maintains some fresh air exchange and limits passenger impact from the outside elements. In some cases, the roof hatches alone may be justified, although it will reduce the airflow inside the bus and the effectiveness of controlling potentially infectious aerosols. The minimal window configurations also allow for bus heaters or HVAC systems to be operated simultaneously to improve comfort levels. However, when it is not possible to open any windows due to weather or to add better air filtration to the buses, consideration should be given to cancelling bus services for that time period due to the increased risks of aerosol transmission.

### Implications for Driver Safety

While the overall study was focused on safety in passenger seating areas, it is important to consider any observations or results that also apply to the safety of bus drivers who spend several hours per day within the bus. There are three situations to describe: the use of auxiliary dashboard fans, the use of driver’s side window for providing fresh air, and airflow dynamics near the driver’s area. In the design of experiments, we considered the supplementary airflows from the dashboard mounted fans to help limit aerosol dispersion to the driver area while also maintaining some airflow toward the rear exhaust windows or roof hatches. Many buses have two such fans installed for windshield defrosting or defogging; one can be aimed toward the back of the bus to aid in the airflow while the other (nearest to the driver) maintains airflow across the windshield. Some buses that have only one such fan, should maintain its use for windshield defogging during certain weather conditions.

During the experiments we observed the NaCl aerosol cloud traveling forward against the small fan’s airflow, however the aerosol peak particle count at the sensor location immediately behind the driver area was significantly attenuated. This per-experiment attenuation at the front particle count sensors compared to the peak particle count nearest the dispersion location in the middle of the bus showed an 85.33% peak reduction on the School bus with windows open and dashboard fans turned on (*n* = 3, particle diameter of 2.5µm) compared to only a 58.23% peak reduction with windows open but without dashboard fans (*n* = 1, particle diameter of 2.5µm). The peak particle count reductions were greater with smaller particle diameters. Adding a third fan to the front of the bus (eleven-inch Honeywell HT-900) reduced the peak by 96.48% near the driver area (*n* = 1, particle diameter of 0.3µm). This suggests that some airflow from the front dashboard fans and driver area can help reduce risk of a driver’s exposure to aerosols sourced from the passenger area. This study, however, did not compare and contrast the effect of dashboard fans on the transit bus experiments.

Regarding the localized airflow effects of having the driver’s side window open, our current analysis does not have sufficient information to provide conclusive results. Future analysis of the data and additional studies should consider detailed monitoring of the airflows and aerosol movement in the driver area with tests that include the driver’s window open and closed, the front passenger door opening, and aerosol dispersion at the front door to simulate a passenger boarding. One concern is if the fans are turned on and the bus windows are closed, they will draw contaminated air from the passenger area and recirculate it, thus increasing driver and passenger risk. In general, the fresh air for the dashboard fans can be provided by the driver’s side window and when the bus door is opened at bus stops, however additional studies should analyze the interior airflow effects with the driver’s window open when the bus is moving while considering the external vehicle airfoil near the driver’s window or other known aerodynamics phenomena. Existing guidance from various transportation agencies also include the recommendation of adding clear plexiglass barriers near the driver for protection from aerosol clouds (Gurley 2020; APTA 2020a) which is expected to alter only the initial aerosol dispersion and to slow the diffusion into the driver’s area. However, future studies should quantify the aerosol diversion offered by the plexiglass barriers. To summarize the implications of this study on driver safety, combining the results of mask effectiveness and the dashboard fans will provide more optimal safety for bus drivers.

### Retrofit Applications to Improve Passenger Air Quality

With regards to modifying the bus HVAC system for improving passenger air quality, there are several strategies and considerations outlined in prior work (Edwards et al. 2021; Stewart et al. 2020). One of the primary strategies is air exchange (ACH) with fresh air which will help evacuate and remove any infectious aerosols (Stewart et al. 2020; Mobile Climate Control 2020; APTA 2020a). Some of the HVAC systems installed in transit buses have a fresh air intake option, while others do not and only have the ability to recirculate air that is exchanged while passenger doors are opened at bus stops. Consequently, a second strategy of improved air filtration must be applied. Another strategy that can also work in parallel is to deactivate infectious diseases using an internal retrofit disinfectant technology such as the UV germicidal irradiation for bus HVAC systems (Huston 2009).

The surface mount application of MERV-13 air filters in this study provided a low-cost and fast method to reduce air contaminants in the bus, however, there are other maintenance-oriented options instead of using removable tape. Some bus HVAC system manufacturers may already provide a mechanism to install MERV-13 pleated filters to balance airflow restriction and HVAC performance. If this is not available, another option is for public transportation fleet service departments to install a simple plastic channel around the transit bus return air grille that can hold the commercial air filters and still allow for servicing. A negative aspect of this retrofit is that without an additional grille cover, the filter media is exposed directly to any passengers near the back seat and could be damaged.

Likewise, the installation of additional small dashboard or ceiling mount fans pointed toward the passenger aisle is a retrofit application that can easily be achieve by public transportation fleet service departments. The commercial vehicle dashboard fans are readily available from local automotive parts suppliers and only require 12-volt standard vehicle power connection. While the installation of an additional fan is achievable, some consideration is needed on costs and installation for a whole fleet of buses.

### Operational Considerations

Since a single strategy will not apply to every situation, public transportation organizations need to consider a layered approach or combinations of strategies to maintain the health and safety of drivers and passengers. Combining the air quality hazard mitigation strategies of remove, replace, divert, deactivate, prevent and protect are all applicable to public transportation (Edwards et al. 2021). One main consideration for public transportation organizations is that the optimal aerosol control configurations and results presented in this study may not be practical in all real-life situations. For example, there are regional differences in weather conditions, school bus pickup and drop off procedures, or longevity of bus routes. Transportation administrators need to consider application of this study’s results and other existing guidelines to their regional challenges by creating policies and training for drivers to safely adapt to daily situations during the COVID-19 pandemic.

Another consideration is to ensure that aerosol risk mitigations are not in conflict with each other. One example is to have only few select windows open for fresh air exchange, such as the “bad weather” test conditions and combine it with HVAC filtration to reduce human respiration emissions. A caution is if the AC compressor were turned on during summer heat conditions while too many windows are open, the evaporator coils can freeze due to the increased workload and possibly cause permanent damage to the HVAC system. In this combined approach, it would be justified to run the HVAC with the AC compressor turned off (depending on the installed HVAC controller system).

### Limitations of This Study

One of the primary limitations of this study is that it does not include results focused on determining safe seating arraignments within the bus. A key observation is that the NaCl aerosol eventually disperses throughout the whole bus, but distinguishment of aerosol cloud exposure levels between individual seats would require instrumentation at every seat (Silcott et al. 2020) rather than a single particle count sensor at every other seat row. Another limitation is that the study is focused on passenger safety rather than understanding localized airflow effects near the driver seat area. A third limitation is that the study considers aerosol dispersion and control from a single simulated cough rather than a real-world scenario with multiple passengers with various respiration rates (inhalation and exhalation). The overall effect on particle count and aerosol dispersion is expected to be more drastic with an increased number of people, however the optimal ventilation schemes would be the same as reported. Determining aerosol exposure risk (time and concentration) with multiple simulated passengers is much more complex and will be considered for future studies or additional mathematical analysis of this data set. A final limitation of this study is that the evaporation rate and aerodynamics of biological aerosols from human exhalation will be slightly different than with the NaCl test agent. Regardless, this study establishes the basis for optimal aerosol dispersion control on the two bus types.

## CONCLUSION

This study presents conclusive results showing the efficacy of masks, open windows, and HVAC systems to reduce risks of aerosol transmission on public transportation buses. The study thoroughly discusses key observations of airflow and aerosol dynamics inside buses and implications or practicalities of applying the results to transportation systems for passenger and driver safety. For example, when masks were not worn and windows are closed, the aerosol particles spread rapidly throughout the whole bus which emphasizes the importance of wearing face coverings and maintaining some airflow to evacuate the particles. In addition, this study also highlights the challenge of an imperfect risk problem where the selected mitigations may not be the optimal solution for all desirable effects such as a ventilation scheme that reduces the overall particle counts but speeds up the dispersion of the aerosol (it can reach more people faster). Considering the tradeoffs between both metrics, the results of this study clearly indicate that ventilation has a greater effect at reducing a passenger’s overall exposure time and concentration to potentially infectious aerosols on the bus. While there are some limitations of this study, it increases scientific understanding of aerosol dispersion and control on buses while bringing clarity on the best options to reduce risks of airborne particle transmission while riding on school and transit buses. While the results and recommendations may seem intuitive, the study provides the scientific data to help inform regional decisions on applying risk mitigation options for COVID-19 variants and other high infectious airborne diseases.

The key recommendations from this study are:

- Require all passengers and drivers to wear masks on buses.
- Open windows partially or fully to make a significant difference in reducing airborne risks.
- Drivers and operators can reduce their risk of exposure by using a dashboard fan to create airflow in the driver seat area.
- In the absence of high fidelity per-seat data, consider seating arrangements that only allow for same household or same cohort passengers to sit together.
- Social distancing of six feet is not practical on most buses, but any extra distance allows the air movement to reduce exposure to infectious particles.
- When it is safe and possible to do so, open transit bus doors at every stop to allow for better air exchange.
- When it is not possible to open windows and doors due to inclement weather, or to add better filtration to the buses, consider cancelling bus services for that time period.

## DATA AVAILABILITY STATEMENT

The datasets generated from this study are available to U.S. national, state and regional agencies as well as healthcare systems on request to the corresponding author.

## AUTHOR CONTRIBUTIONS

N.E., R.P., M.G., and R.W. designed the research; N.E., R.P., M.G., R.W., J.W., J.S, and B.B. performed research; N.E., M.G., R.W., J.W., J.S, and A.E.C. analyzed data; N.E., R.P., J.W., and J.S. wrote the paper.

## ACKNOWLEDGMENT

Our team acknowledges Colorado Springs School District 11 and City of Colorado Springs Mountain Metro for providing use of their buses along with a driver, and invaluable conversations on operational implications of the field results.

## FUNDING/SUPPORT

This work was supported by independent research and development funding provided by the author’s organization, The MITRE Corporation. ©2021 The MITRE Corporation. ALL RIGHTS RESERVED. Approved for Public Release; Distribution Unlimited. Public Release Case Number 21-0157.

## Supplementary Information

**Figure S1:**
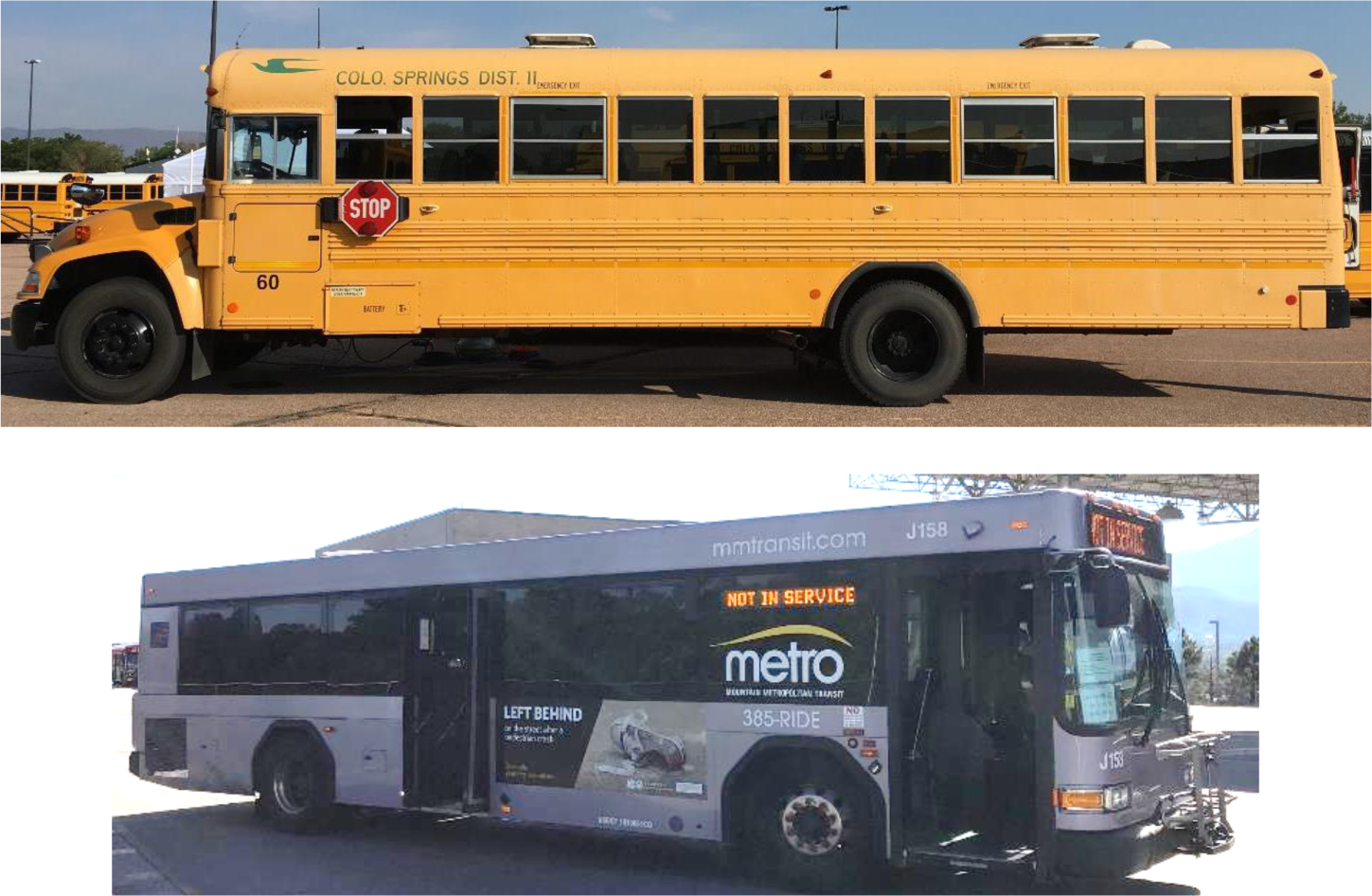
Buses used for the study. (A) 66 seat 2013 Blue Bird Vision propane school bus. (B) 35 foot 2015 Gillig G27B low-floor transit bus.

**Figure S2:**
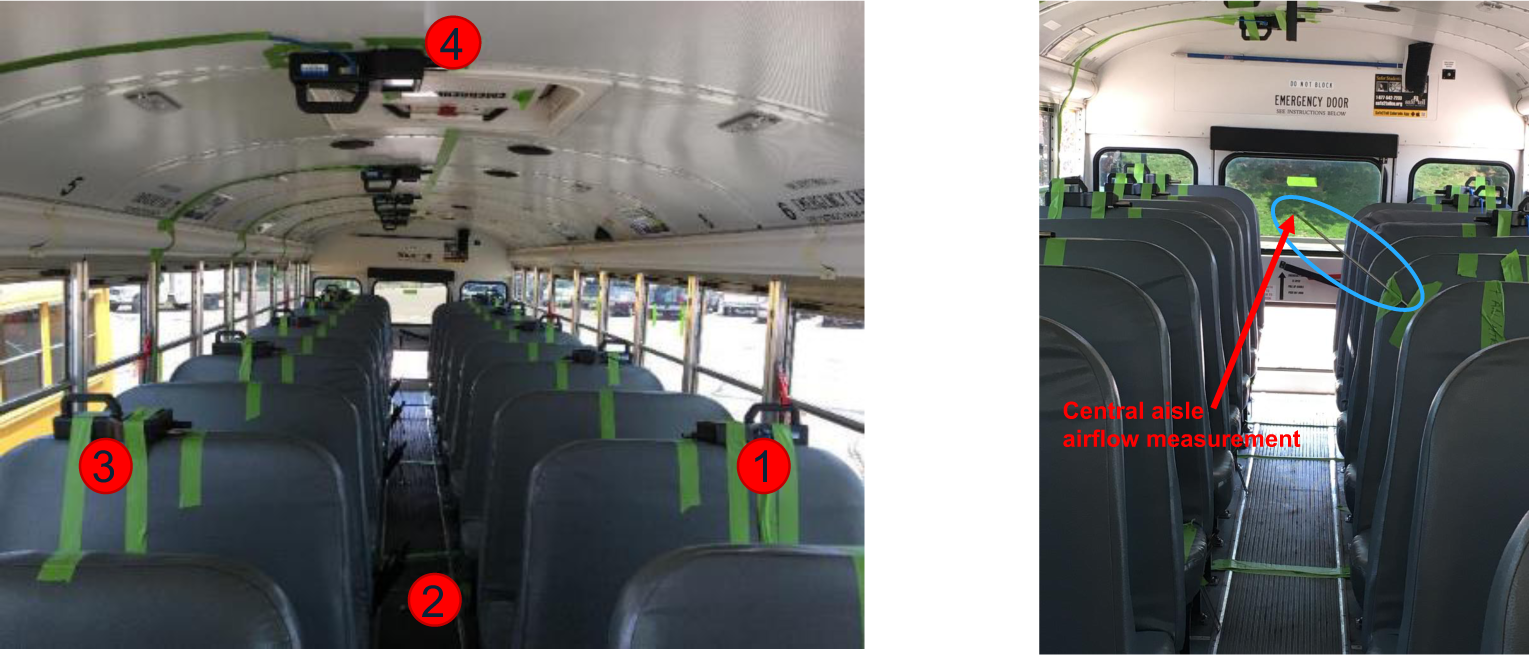
Layout of the school bus sensors: floor, ceiling, and passenger seats. Image on right is showing the mounted location of the anemometer for measuring central airflow.

**Figure S3:**
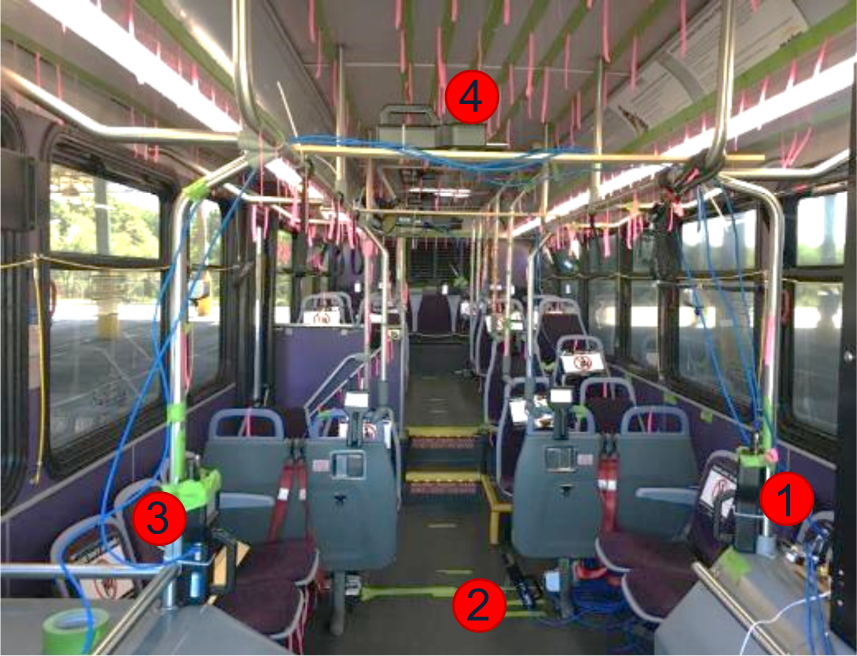
Layout of the transit bus sensors: floor, ceiling, and passenger seats.

**Figure S4:**
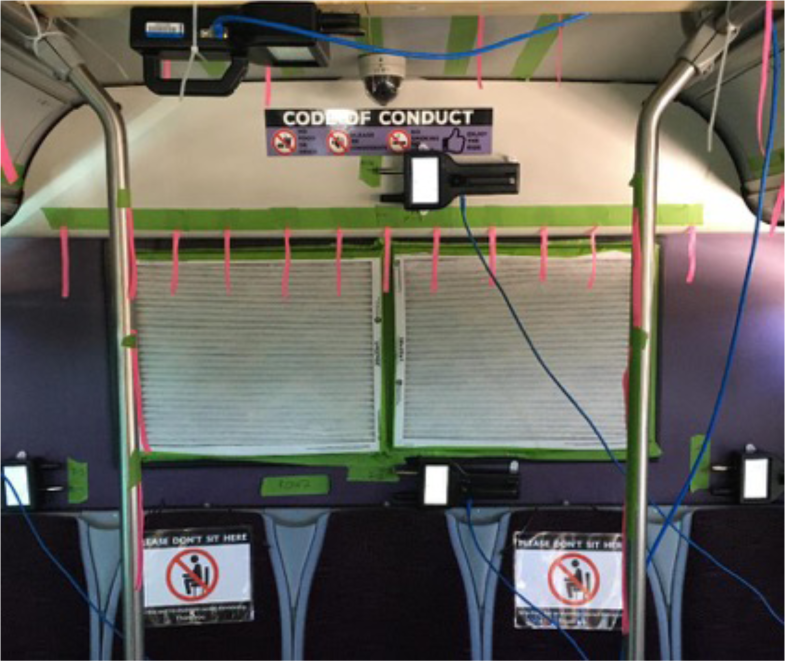
Application of 2 quantity MERV-13 air filters to the transit bus HVAC return air grille in the back of the bus.

**Figure S5:**
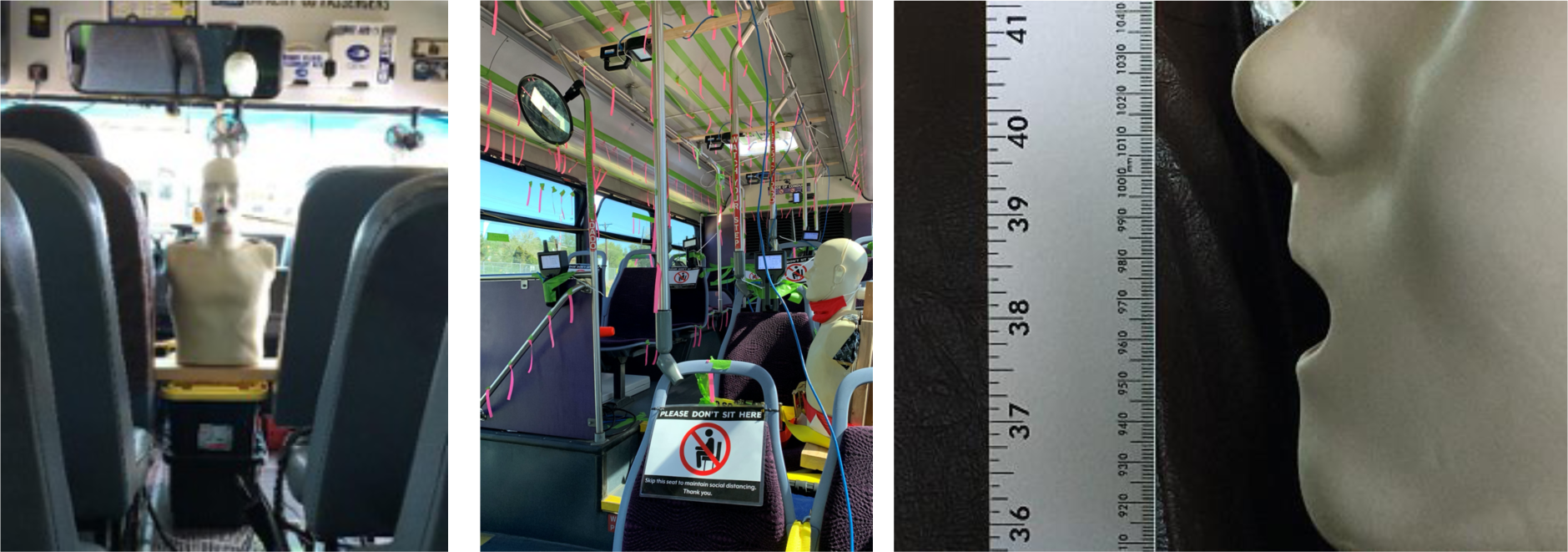
Exhalation simulator shown (A) in the middle aisle of the school bus, (B) in the middle seat location of the transit bus, and (C) the dispersion height of 365 cm from the floor.

**Figure S6:**
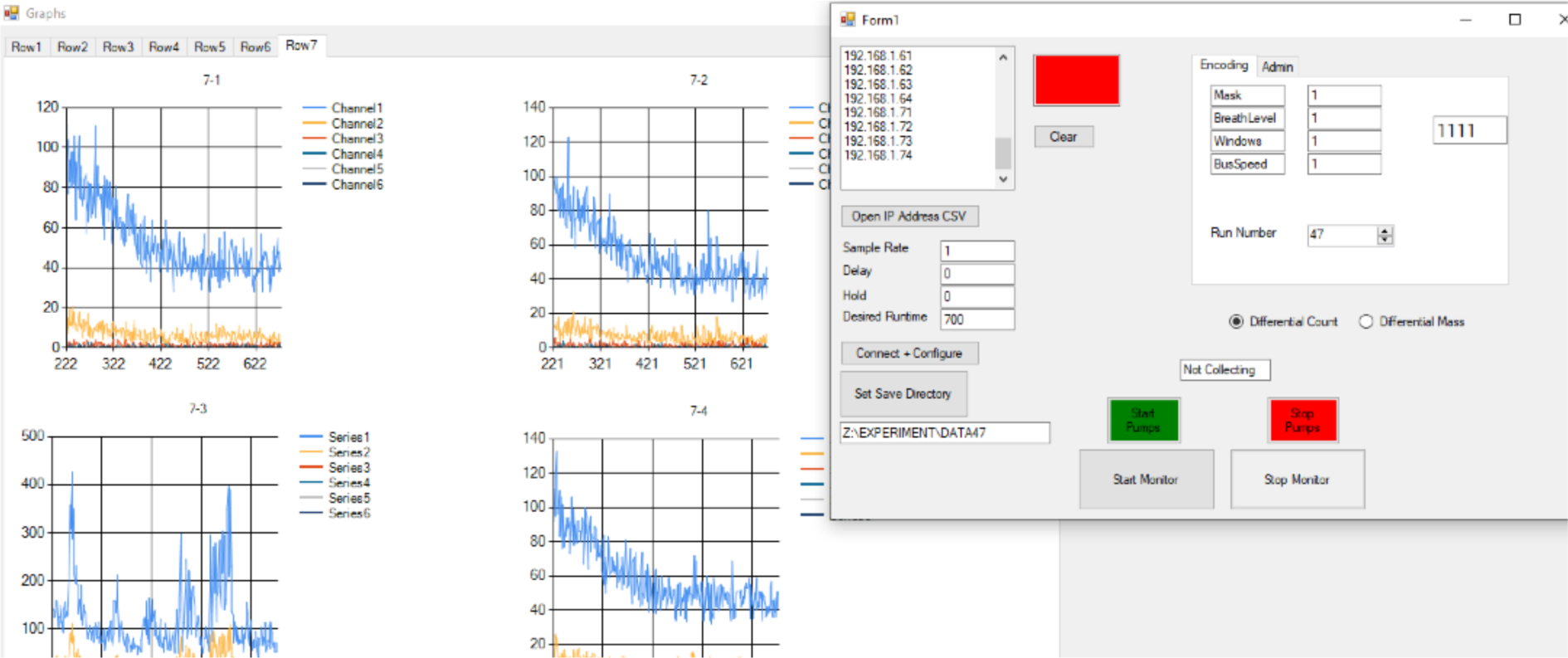
Screenshot of the custom multithreaded supervisory control and data acquisition (SCADA) software for nanoparticle dispersion test equipment

**Figure S7:**
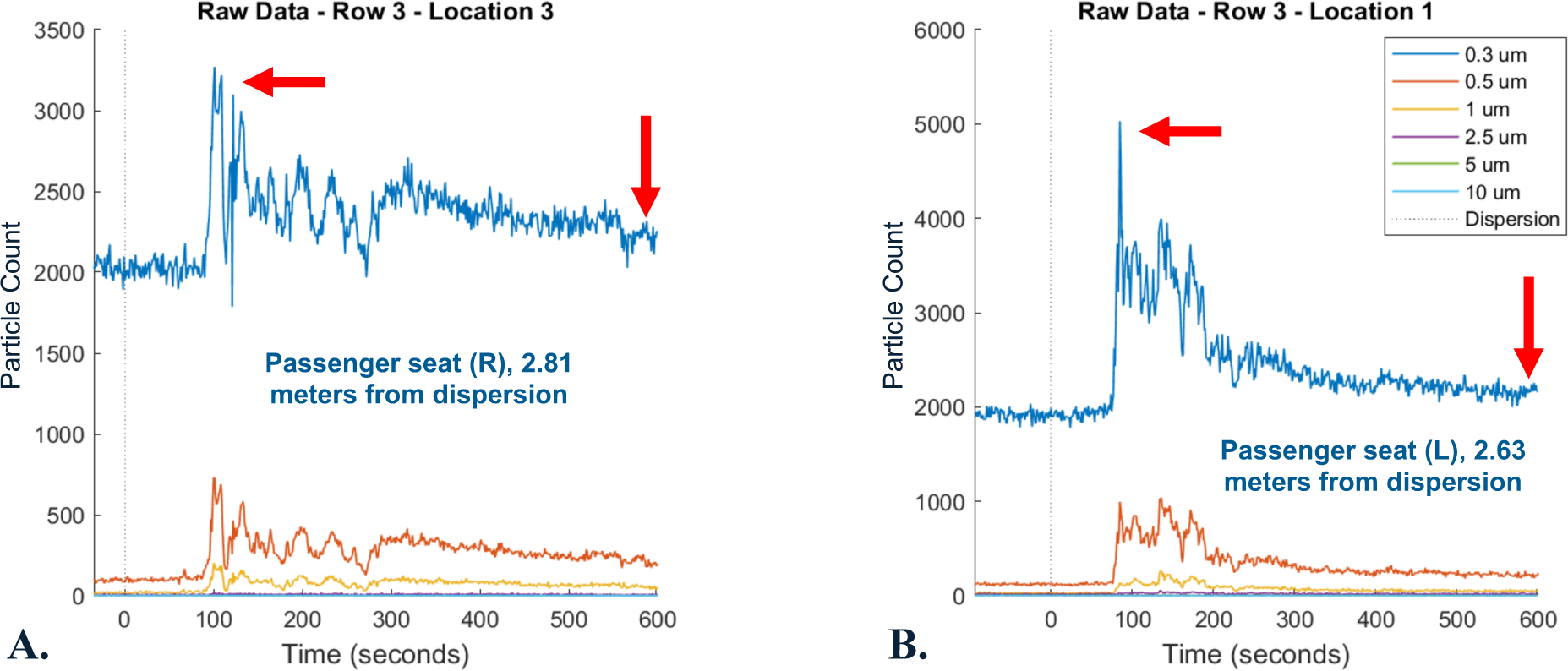
Particle count time-series waveforms on the school bus (A) and transit bus (B) showing the effect of a mask which reduces the overall particle counts (left arrow and AUC) while elongating the residence of particles (right arrow)

**Figure S8:**
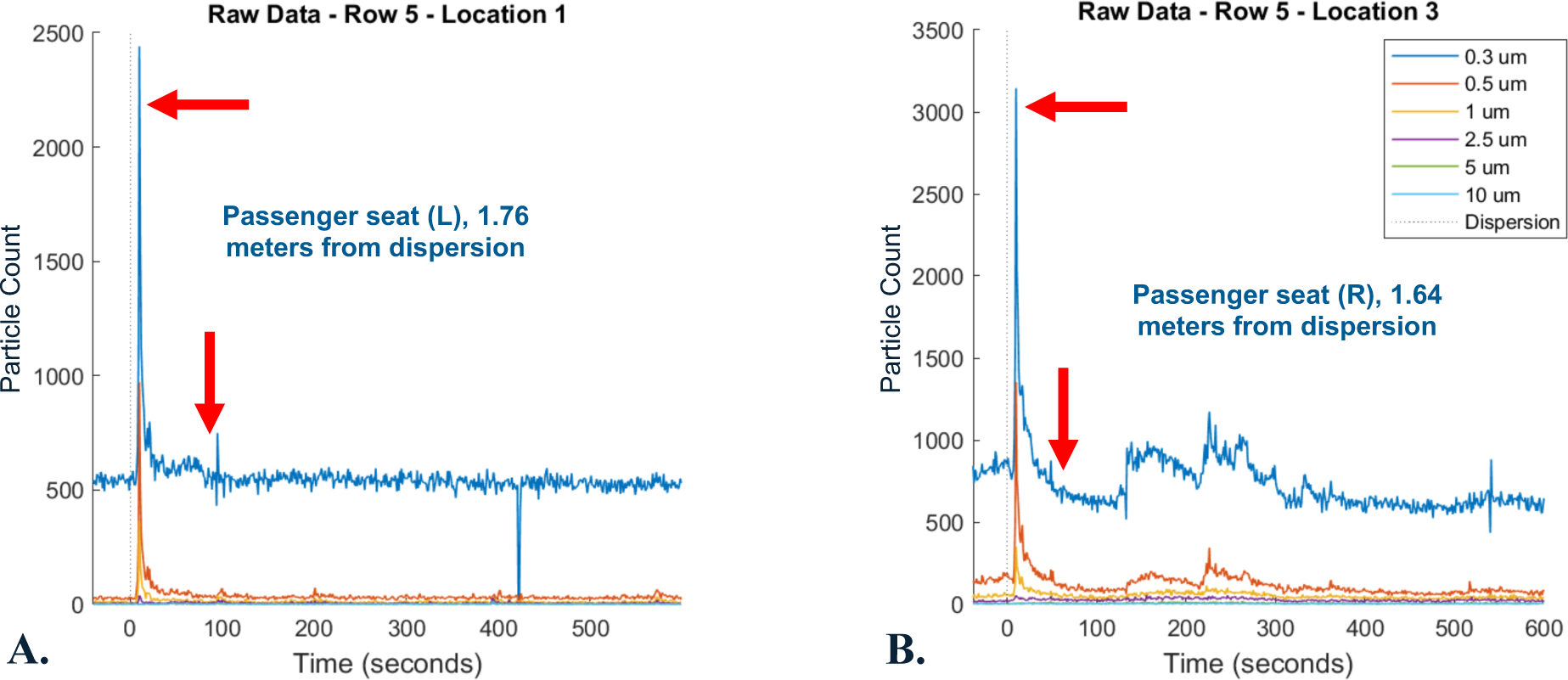
Particle count time-series waveforms on the school bus (A) and transit bus (B) showing the effects from all windows, fans, and roof hatches with a narrow peak (upper arrow) and a short residence time of particles (lower arrow).

**Figure S9:**
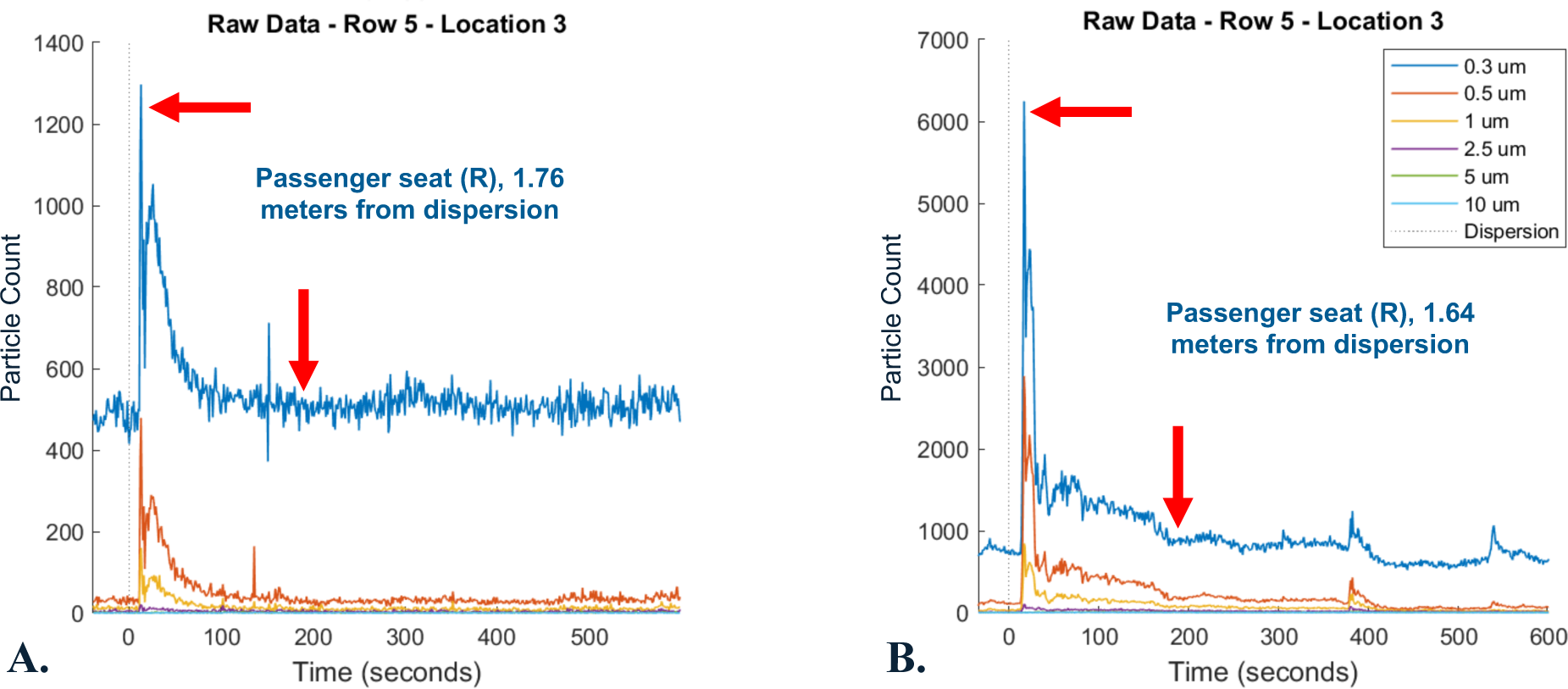
Particle count time-series waveforms on the school bus (A) and transit bus (B) showing the effects from the inclement weather configurations with a slightly widened peak (upper arrow) and slightly elongated residence time of particles (lower arrow) compared to all windows open.

**Figure S10:**
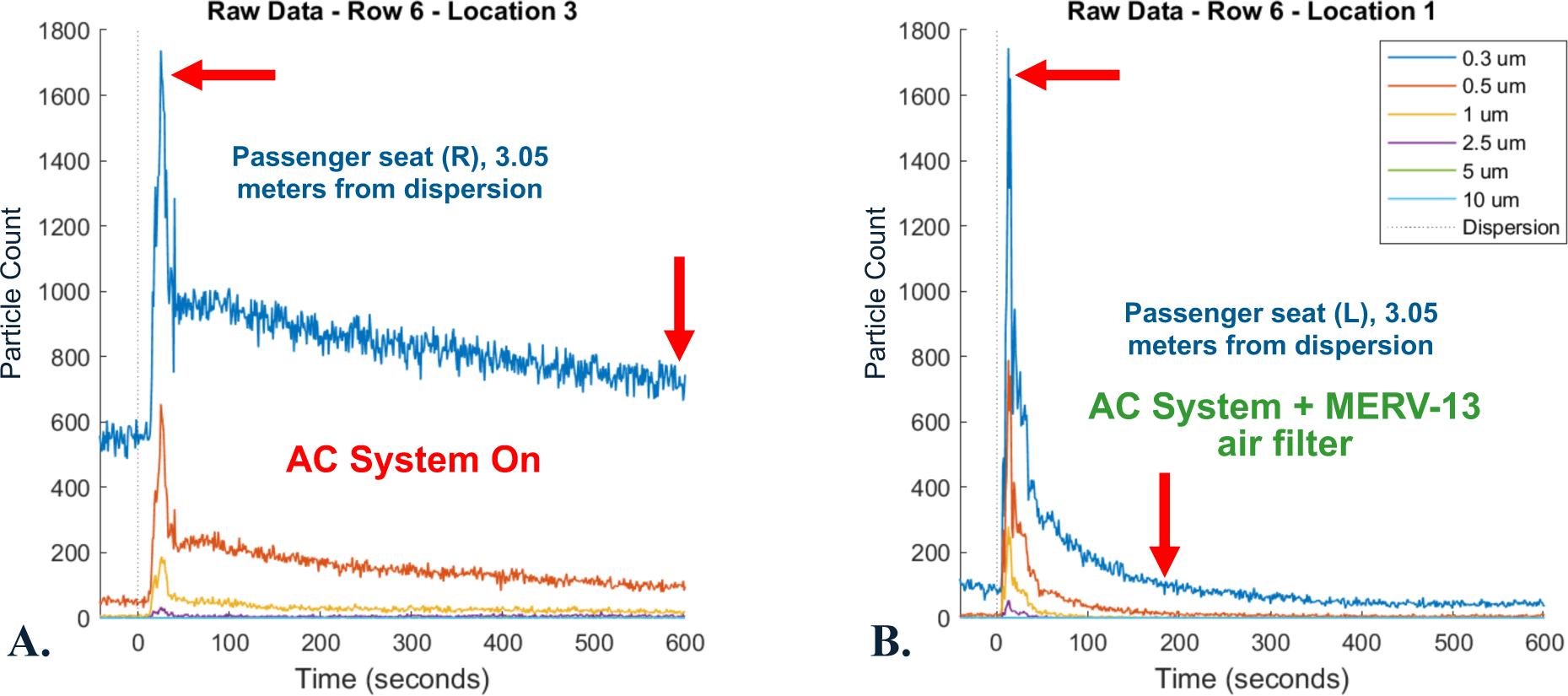
Particle count time-series waveforms from the transit bus HVAC system showing the effect when the AC system is turned on (A) with a slightly widened peak (upper arrow) and a gradual decrease of particles but never reaching a nominal level (lower arrow). When a MERV-13 air filter is applied (B), the peak is also slightly widened but the decrease of particles is more rapid (residence time of particles < 4 min).

**Figure S11:**
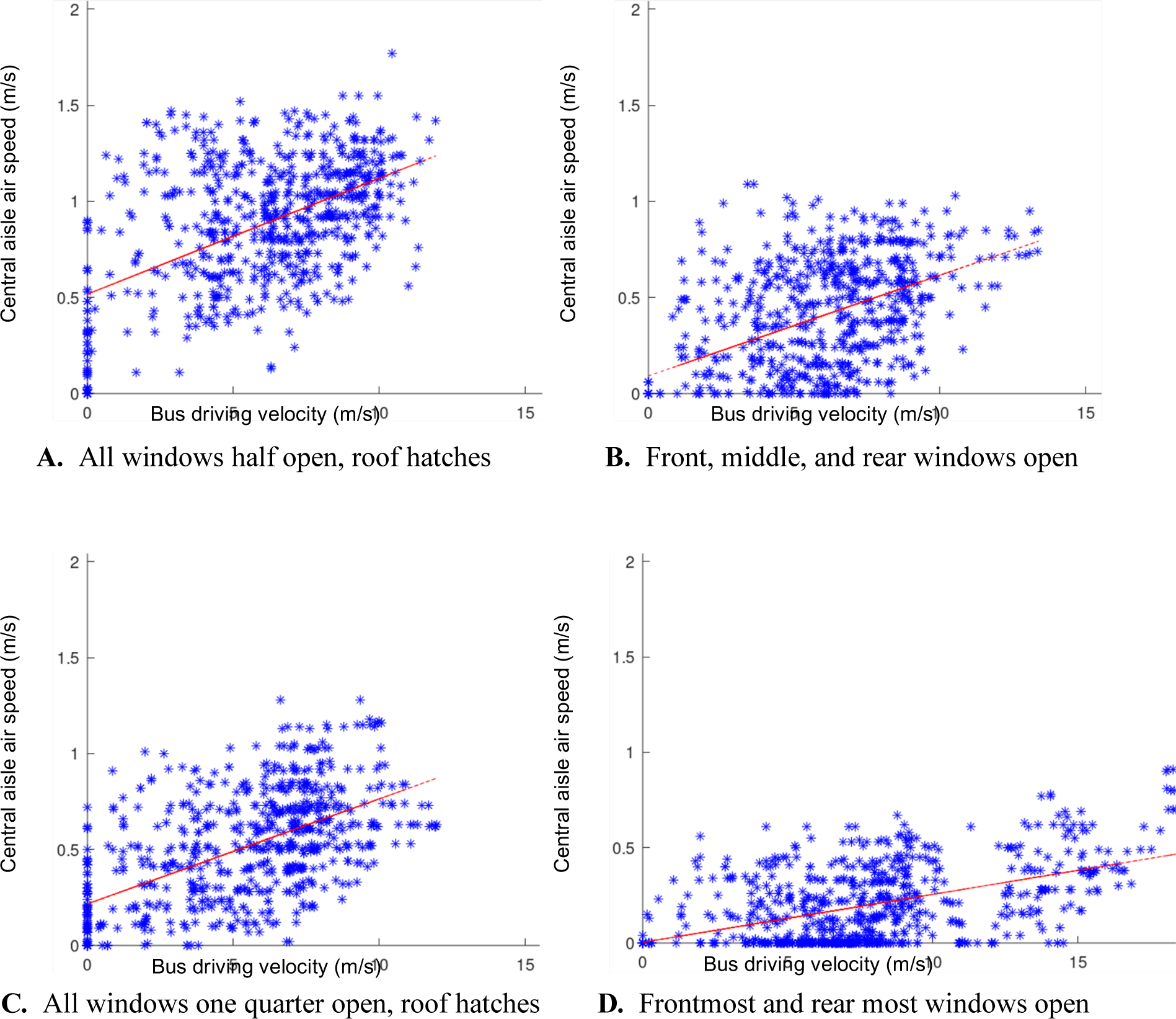
Airflow compared to school bus velocity. Anemometer was placed in a central location of passenger area aisle. Red line shows the approximate linear fit.

**Figure S12:**
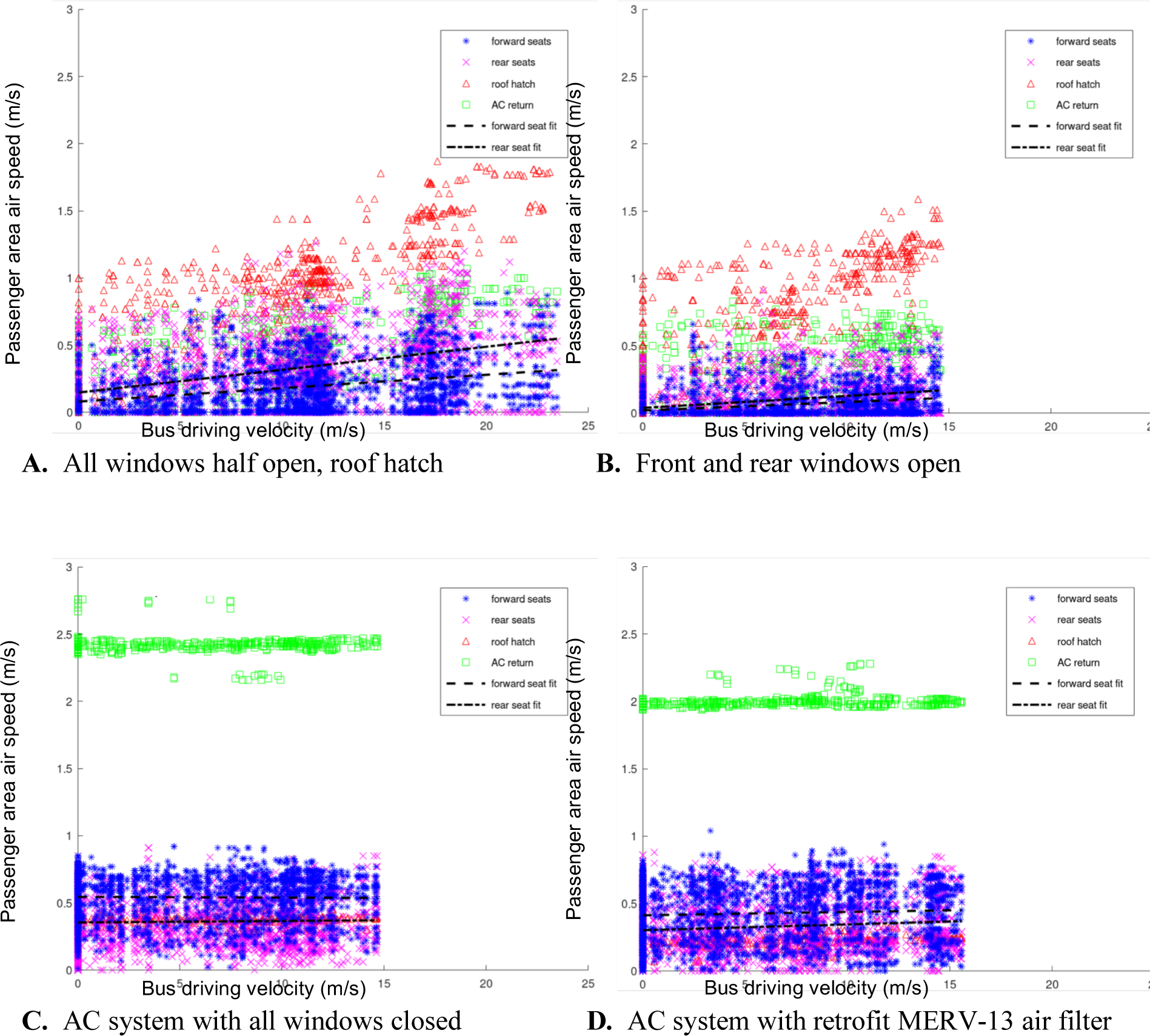
Airflow compared to transit bus velocity. 12 Anemometers placed at different sensor locations throughout the passenger area.

**Figure S13:**
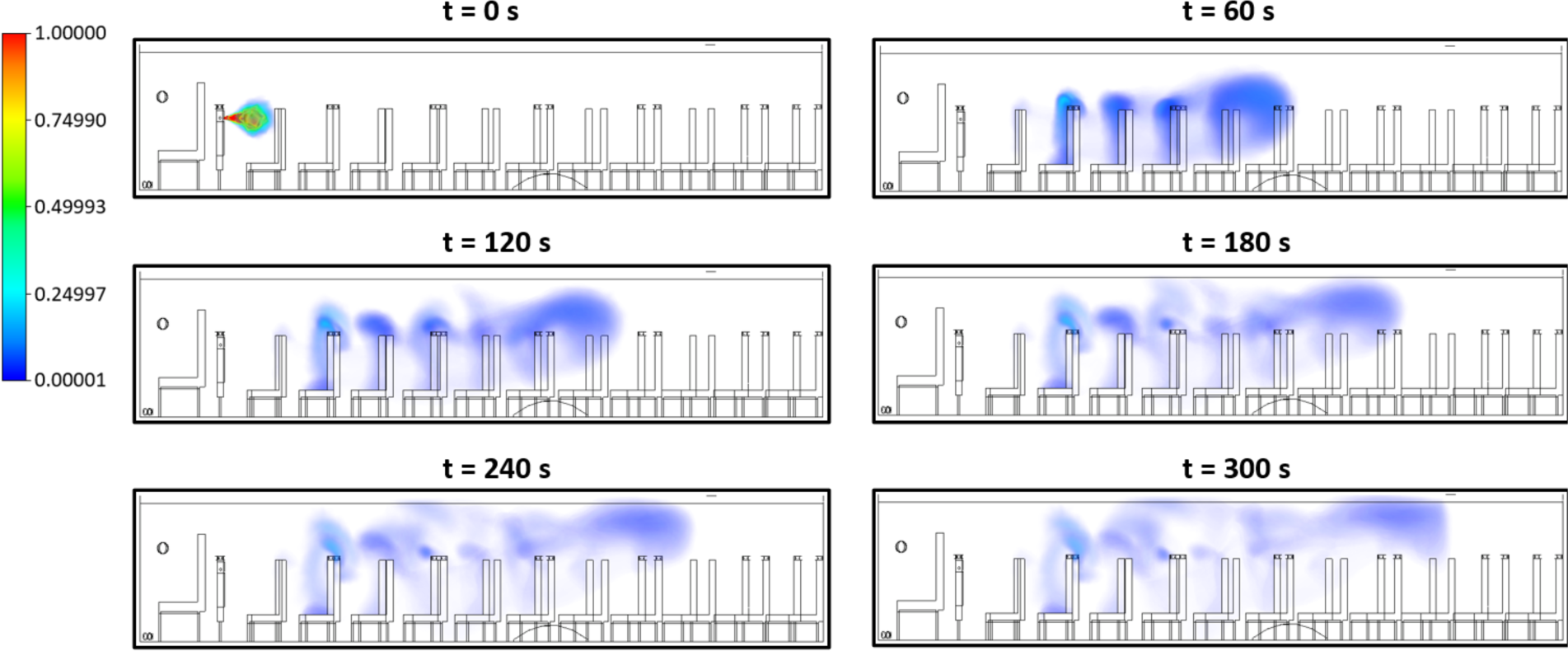
CFD simulation of aerosol dispersion on the school bus showing a baffle effect from the high back seats where regions of high aerosol concentration are present at the front side of seats causing more rapid diffusion of the initial aerosol jet.

**Table S1:** Comparison of 2019 and 2020 quarterly bus ridership using estimated unlinked transit passenger trip data.

| Estimated Unlinked Transit Passenger Trip |  |  |  |  |  |
| --- | --- | --- | --- | --- | --- |
| Year | Q1 | Q2 | Q3 | Q4 | Annual Total |
| 2019 | 1140.05 M | 1180.32 M | 1169.05 M | 1175.33 M | 4664.75 M |
| 2020 | 1026.92 M | 409.93 M | 566.54 M | Not available | -- |

Source. APTA. 2020. "Ridership Report." American Public Transportation Association. https://www.apta.com/research-technical-resources/transit-statistics/ridership-report/.

**Table S2:**
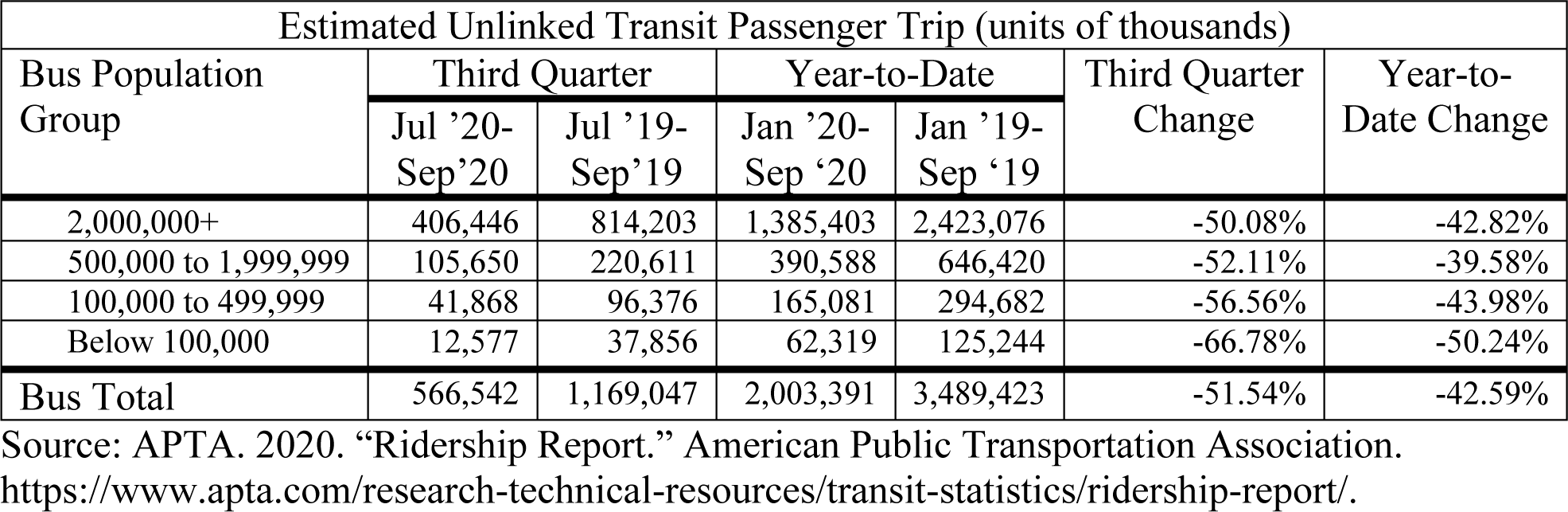
Comparison of community population demographics of Q3, 2020 bus ridership.

**Table S3:**
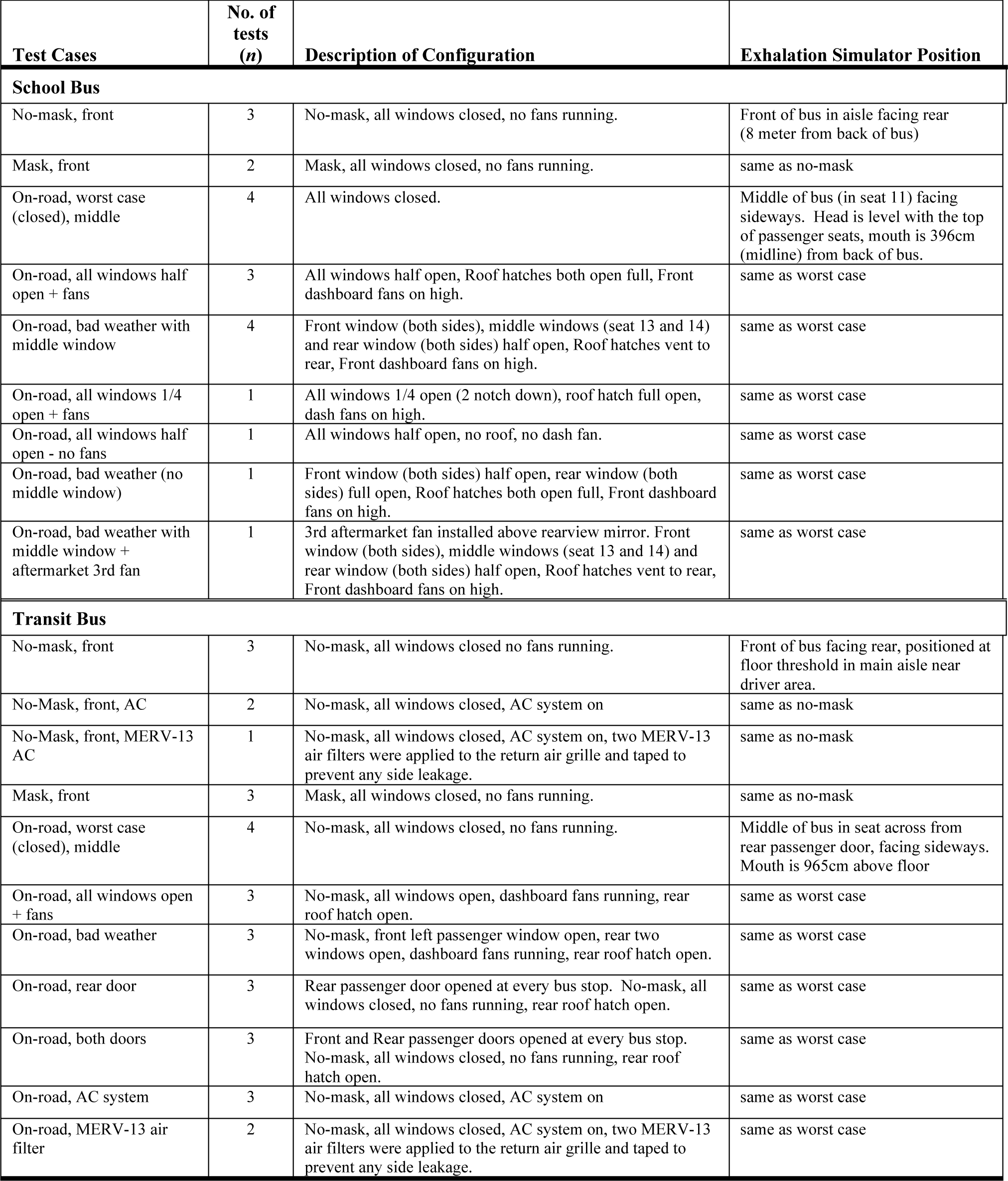
Test Configurations of the School Bus and Transit Bus

**Table S4:** Linear distances for particle counter sensors

| **Sensor Row** | **Dist from Front**<br>**Dispersion (meter)** | **Dist from Middle**<br>**Dispersion (meter)** |
| --- | --- | --- |
| **School Bus** |  |  |
| Row 1 | 0.00 | 4.02 |
| Row 2 | 1.36 | 2.66 |
| Row 3 | 2.81 | 1.21 |
| Row 4 | 4.35 | -0.33 |
| Row 5 | 5.78 | -1.76 |
| Row 6 | 7.24 | -3.22 |
| Row 7 | 7.86 | -3.84 |
| **Transit Bus** |  |  |
| Row 1 | 0.00 | 4.35 |
| Row 2 | 0.89 | 3.46 |
| Row 3 | 2.63 | 1.72 |
| Row 4 | 4.50 | -0.15 |
| Row 5 | 5.98 | -1.64 |
| Row 6 | 7.40 | -3.05 |
| Row 7 | 8.29 | -3.95 |

## Notes

### Competing Interest Statement

The authors have declared no competing interest.

### Author Declarations

The MITRE Institutional Review Board reviewed this study protocol, Quantifying Airborne Dispersion in Public Transportation, and determined that the activity is not human subjects research as defined by 45 CFR 46.

